# COVID-19 vaccine booster strategies in light of emerging viral variants: Frequency, timing, and target groups

**DOI:** 10.1101/2022.06.22.22276760

**Authors:** Sherrie L Kelly, Epke A Le Rutte, Maximilian Richter, Melissa A Penny, Andrew J Shattock

## Abstract

**Background:** Vaccinations have reduced severe burden of COVID-19 and allowed for lifting of non-pharmaceutical interventions. However, with immunity waning alongside emergence of more transmissible variants of concern, vaccination strategies must be examined.

**Methods:** Here we apply a SARS-CoV-2 transmission model to identify preferred frequency, timing, and target groups for vaccine boosters to minimise public health burden and health systems risk. We estimated new infections and hospital admissions averted over two-years through annual or biannual boosting of those eligible (those who received doses one and two) who are 1) most vulnerable (60+ or persons with comorbidities) or 2) those 5+, at universal (98% of eligible) or lower coverage (85% of those 50+ or with comorbidities and 50% of 5−49-year-olds who are eligible) representing moderate vaccine fatigue and/or hesitancy. We simulated three emerging variant scenarios: 1) no new variants; 2) 25% more infectious and immune-evading, Omicron-level severity, variants emerge annually and become dominant; and 3) emerge biannually. We further explored the impact of varying seasonality, variant severity, timing, immune evasion, and infectivity, and vaccine infection blocking assumptions.

**Results:** To minimise COVID-19-related hospitalisations over the next two years, boosters should be provided for all those eligible annually three-four months ahead of peak winter whether or not new variants of concern emerge. Only boosting those most vulnerable is unlikely to ensure reduced stress on health systems. Moreover, boosting all eligible protects those most vulnerable more than only boosting the vulnerable group. Conversely, more hospitalisations could be averted per booster dose through annual boosting of those most vulnerable versus all eligible, an indication of cost-effectiveness. Whereas increasing to biannual boosting showed diminishing returns. Results were robust when key model parameters were varied. However, we found that the more frequently variants emerge, the less the effect boosters will have, regardless of whether administered annually or biannually.

**Conclusions:** Well-timed and targeted vaccine boosters preferencing vulnerable, and if possible, all those eligible to receive boosters, can minimise infections and hospital admissions. Findings provide model-based evidence for decision-makers to plan for administering COVID-19 boosters ahead of winter 2022−2023 to help mitigate the health burden and health system stress.

## Background

Between December 2019 and May 2022, 529 million confirmed cases of COVID-19 and over 6.29 million COVID-19-related deaths were reported.^1^ As of May 2022, over 11.9 billion COVID-19 vaccine doses have been administered worldwide, but as SARS-CoV-2 variants continue to spread, increasing global vaccine coverage will be crucial. Rollout strategies for first and second vaccine doses are being combined with subsequent booster doses to bolster immunity levels. Rapid viral evolution and the relatively short period that COVID-19 vaccines have been available have limited our ability to observe vaccine efficacy and duration over long periods. For example, a study conducted in England from the end of November to early December 2021 reported vaccine effectiveness against symptomatic disease from the Omicron variant two to nine weeks following the second dose of the Pfizer/BioNTech mRNA vaccine to be relatively high, at approximately 88%. While this protection was reported to have waned to roughly 65% after 20 to 24 weeks, a booster dose was reported to boost protection to approximately 92% after two weeks.^2^ This is reassuring for those who received their booster dose and are thus benefiting from a lower probability of becoming infected and developing COVID-19-related disease, but highlights the need for a longer-term vaccination strategy that accounts for waning immunity, particularly with the likelihood of the emergence of new immune-evading variants of concern.

Prior to Omicron subvariants, new SARS-CoV-2 variants had emerged and become dominant about every six months,^3^ and while the overall risk of severe disease or death from Omicron infection is lower than from the previously dominant Delta variant, Omicron subvariants are more infectious and their rapid spread is still causing health system stress.^4^ Long-COVID may also continue to contribute to the public health burden. Furthermore, variants of the Omicron lineage are continuing to evolve and make gains by eroding host immunity. If this continues, then SARS-CoV-2 epidemics may reach steady state and become endemic similar to influenza. In this case, however, when population-wide immunity declines, circulating variants with immune-evading mutations could become dominant, may potentially be more severe and periodic waves may arise.^3^ This emphasises the urgency for everyone to receive at least one COVID-19 vaccine and highlights the need for strategic delivery of vaccine booster doses.

Using our individual-based SARS-CoV-2 transmission model, OpenCOVID, we simulated a variety of vaccination strategies and model assumptions, to identify the preferred frequency, timing, and target population for future vaccine booster doses in light of potential emerging viral variants. Model outcomes are used to assess the impact of different booster strategies on public health outcomes and health system risks.

## Methods

### Model framework

Our individual-based model, OpenCOVID, is a stochastic discrete-time model of SARS-CoV-2 transmission and COVID-19 disease progression and response.^5^ Our individual-based model, OpenCOVID, is a dynamic stochastic discrete-time model of SARS-CoV-2 transmission and COVID-19 disease progression and response.^6^ The model simulates viral transmission between infectious and susceptible individuals that come in contact through an age-structured, small-world network. The probability of transmission in each exposure stage is influenced by the infectiousness of the infected individual, the immunity of the susceptible individual (acquired through previous infection and/or through vaccination), and a background seasonality pattern (reflecting a larger proportion of contacts being in closer contact indoors as the temperatures became cooler). Infectiousness is a function of variant infectivity and time since infection. Once infected, a latency period is followed by a pre-symptomatic stage, after which an individual can experience asymptomatic, mild, or severe disease. Severe cases can lead to hospitalisation, intensive care unit admission, and ultimately death (see Supplementary Table S1 for further details on prognosis probabilities by age, following infection with SARS-CoV-2 Omicron variant). Recovery after infection leads to development of immunity. This immunity is assumed to wane over time as illustrated in Figure S1, with the risk of new infection depending on the probability of exposure and properties of existing and potential novel SARS-COV-2 variants (i.e., infectiousness and immune evading profile). Briefly, the model captures immunity decay for naturally acquired and vaccine induced infection and seasonality (as illustrated in Supplementary Information Figure S1 and S2, respectively), infection states, non-pharmaceutical interventions (NPIs) can be included (as further described in the *Methods* section *Non-pharmaceutical interventions)*, can be tailored to a specific setting or as investigated here an archetypal setting with Re fitted to 1.0 in spring (as further described in the *Methods* section *Model parameterisation and initialisation)* in the absence of any NPIs assuming immunity from natural infection and vaccination is similar to profiles for Europe in early 2022 during the first Omicron wave). A detailed description of the OpenCOVID model is provided in ^6, 7^ with model code publicly available at https://github.com/SwissTPH/OpenCOVID.

### Model parameterisation and initialisation

We simulate an 850-day period, approximately two years and four months from spring in year one to summer in year three. We apply a global demographic distribution^8^ to a simulated population of 100,000 people, and assume 40% of the population had previously been infected with SARS-CoV-2 at least once over the two-year simulation period.

The model was fitted to allow an archetypal setting with an effective reproduction number, R_e_(r), of 1.0 at the start of the simulation period, representing the shoulder season between winter and summer in year one. This represents the global trend for case numbers when the Omicron variant emerged and became the dominant variant in late 2021. Using an average number of daily contacts the model was fitted to this R_e_(r) of 1.0 in early spring 2022. This inherently captures the effect of any non-pharmaceutical interventions that were in place at that time. To reflect the element of chance that naturally occurs in model transmission dynamics, 100 random stochastic simulations were performed for each scenario with 95% prediction intervals presented.

### Seasonality

Simulations represent an archetypal setting approximately representative of Northern hemisphere seasonality. Seasonality is assumed to follow a scaled cosine function. It oscillates between minimum seasonal forcing during the peak summer period, and maximum forcing during peak winter months as illustrated in Supplementary Information Figure S2. As people tend to remain indoors during the winter, the probability of transmission per contact is increased during winter months. The best estimate for the seasonal scaling factor was taken from a previous study assessing the COVID-19 epidemic in Switzerland.^7^ This previous study simultaneously fit this seasonal scaling factor parameter along with several other parameters to align the OpenCOVID model to epidemiological data. To assess the effect of the seasonal scaling factor on model outcomes, this value was subjected to a sensitivity analysis.

### Non-pharmaceutical interventions

To examine the sole effect of vaccinations on the spread of SARS-CoV-2 and COVID-19 disease progression, we did not explicitly model any non-pharmaceutical interventions (NPIs) such as physical distancing and facemask usage and lockdowns in future scenarios. However, since a proportion of the population will likely continue to wear masks in some situations such as on public transport in certain settings, this protective effect was captured indirectly by using the number of effective network contacts to calibrate the model to the effective reproduction number at the start of the simulation period.

### Vaccination strategy

In this analysis, we simulate the impact of the first-generation mRNA vaccine (here we modelled vaccine profile for the Pfizer/BioNTech (BNT162b2)), which was developed with specificity to the dominant SARS-CoV-2 variant at the time, the Alpha variant. Vaccines have a two-fold effect; first, they provide protection against new infection through development of immunity. Secondly, once infected, vaccines reduce the probability of developing severe symptoms, leading to reduced hospitalisations, intensive care unit admissions, and potentially death.

Individuals who previously received primary vaccination (doses one and two) prior to the start of the simulation period were considered eligible to receive booster doses. Default universal coverage of booster doses was set to 98% coverage of those eligible. We assume 1,500 booster doses per 100,000 people per day (1.5% of the simulated population) could be administered, based on achievable vaccine delivery rates across several countries.^9^ Doses were administered sequentially by descending age and comorbidity risk group as described in the *Scenario design* section of the *Methods*.

Following administration of each booster dose, vaccine-induced immunity is assumed to immediately peak at 85%^10^ before exponentially waning to 15% with a half-life of 105 days^5^ (see Supplementary Information Figure S1). The infection-blocking component of the vaccine was assumed to represent 80% of the overall 85% vaccine efficacy, with the remaining 5% attributed to preventing infections from progressing to severe disease. This infection-blocking value was subjected to a sensitivity analysis. Vaccine protection is modelled in the context of naturally acquired immunity following SARS-CoV-2 infection, whereby natural immunity is assumed to reach peak immunity of 95% aligned with findings from Chivese and colleagues,^11^ before waning exponentially to 20% in 600 days.^12^

### Variant properties

At the time of writing, Omicron was the dominant SARS-CoV-2 variant worldwide, with subvariants BA.1, BA.2, BA.4, BA.5, and BA.2.12.1 having emerged along the Omicron lineage, having replaced the previously dominant Delta variant (B.1.617.2). Based on previous global trends, we assume conservatively that new variants of concern will emerge ahead of the winter season at six or twelve month intervals. We assume a variant profile with 25% higher infectivity than the previously dominant variant (a transmission multiplication factor of 1.25 per exposure), with immune evasion properties of 25%, but with the same severity as the Omicron variant. The probability that immunologically naïve individuals who become infected will develop severe disease is dependent on variant severity, but also the individuals’ age and comorbidity status, with further details described in ^7^. To assess the effect of variant properties on model outcomes, values for variant infectivity, severity, and immune-evading capacity were subjected to a sensitivity analysis.

### Scenario design

Scenarios were designed to model the impact of vaccine booster doses administered at different frequencies, coverage levels, timings, and to different target groups. A baseline scenario was established whereby the primary series vaccination (doses one and two) was implemented prior to the simulation period for those five years and older, with no booster doses given during the simulations. Baseline vaccination coverage is 93% of those most vulnerable (those 60 years and older following WHO reports^13^ or persons with comorbidities), 90% of 50−59-year-olds, 80% of 30−49-year-olds, and 75% of 5−29-year-olds.

All other scenarios are identical to baseline with primary vaccination (doses one and two) administered pre-simulation at baseline coverage, but with first-generation booster doses (three and up) administered during the simulation to 98% of those who previously received the primary series, that is those eligible to receive booster doses, with this referred to as universal booster coverage. Vaccine boosters were administered every six or twelve months, starting in year one ahead of summer for the biannual booster scenario or ahead of winter for the annual scenario. For all booster scenarios, administering doses to those most vulnerable every six months also includes boosting all eligible every twelve months. Lower booster coverage of those eligible, was administered first to 85% of those most vulnerable (those 60 years and older or persons with comorbidities), then 85% of individuals 50−59 years of age, followed by 50% of 30−49-year-olds, and lastly 50% of 5−29-year-olds. Lower booster coverage is an approximate reflection of booster coverage levels in Europe at the time of writing and represents moderate vaccine fatigue and/or hesitancy.^9^

### Sensitivity analysis

A sensitivity analysis was conducted to assess the impact of varying key model parameter inputs, as detailed in Table 1, on projected outcomes.

**Table 1:** Inputs used for the sensitivity analysis of key parameters.

| Parameter | Parameter effect | Best estimate | Lower bound | Upper bound |
| --- | --- | --- | --- | --- |
| Seasonality-scaling factor (dimensionless)* | Strength of seasonal forcing on infectiousness per exposure | 0.30<sup>6</sup> | 0.20 | 0.40 |
| Emerging variant severity (dimensionless) | Multiplicative probability of severe disease for each new emerging variant | 1.00 | 0.80 | 1.20 |
| Emerging variant timing | Impact of when novel variants emerge and become dominant | 3.5 months before peak winter | -1 month | +1 month |
| Vaccine infection-blocking (%) | Infection-blocking effect of vaccination (remainder is severe disease prevention) | 0.85<sup>†</sup> | 0.60 | 0.95 |
| Emerging variant immune evasion (%) | Impact of immune evasion on variant infectiousness | 0.25 | 0 | 0.50 |
| Emerging variant infectivity (ratio) | Multiplicative probability of transmission per exposure for each emerging variant | 1.25 | 1.10 | 1.40 |

\*See Supplementary Information Figure S2 for the seasonality profile.
<sup>†</sup>As shown in Supplementary Information Figure S1.
No effect was observed from varying daily vaccine booster capacity (results not shown) from 1,500 (best estimate) to 750 doses (lower bound) and 2,250 (upper bound) per day in a simulated population of 100,000 people following default timing to start administering doses.

## Results

We examined the benefit of targeting booster doses to those eligible to receive boosters who are either most vulnerable (those 60 years and older or persons with comorbidities) or all those eligible, five years and older using the described age and risk factor priority scheme, in settings with either no new SARS-CoV-2 variants emerging or with new variants 25% more infectious and immune-evading, but with the same level of severity as the Omicron variant, emerging annually. We found that, the biggest relative impact was from annual boosting of all those eligible, which was consistent with or without new emerging variants (Figure 1, blue dashed and solid curves, respectively), given the assumptions modelled for waning immunity. In the case where no new variants emerge, boosting only those most vulnerable every twelve months led to a 32% (95% CI 24−40%) reduction in hospital admissions over a two-year period compared with no boosters. If annual boosting were to be expanded to those eligible, a 70% (95% CI 65−73%) reduction in hospitalisations could be achieved over this period. If boosting frequency were increased to every six months and targeted to only those most vulnerable (while continuing to boost those eligible annually), then a 77% (95% CI 74−80%) reduction could be realised, an over two-fold decrease in admissions. Extending biannual boosting to those eligible is projected to have diminishing returns, with an 81% (95% CI 79−83%) reduction in hospitalisations over this period. Keeping in mind that the numbers of booster doses required per 100,000 people per year for these four strategies (annual boosting of most vulnerable, eligible annually, vulnerable biannually with eligible annually, and eligible biannually) is 26,000, 78,000, 104,000, and 156,000, respectively, with those most vulnerable representing a third of those eligible to receive boosters.

**Figure 1:**
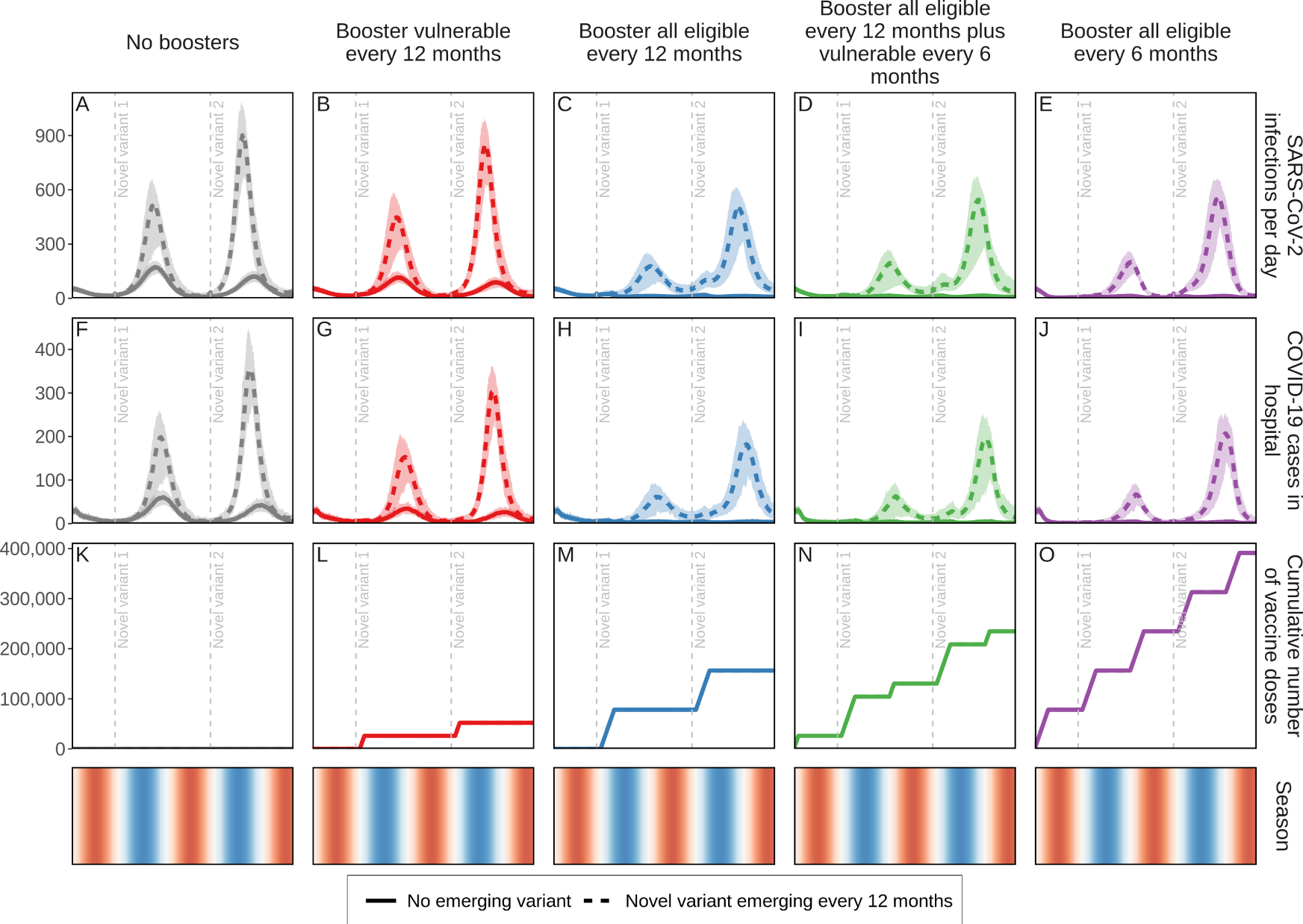
Daily projected impact on SARS-CoV-2 infections and COVID-19-related hospital admissions in just over a two-year period in a population of 100,000 individuals. Those who previously received vaccine doses one and two were eligible to receive a first-generation COVID-19 vaccine booster dose. Individuals who received boosters every twelve or six months are aggregated in two groups, (1) those most vulnerable, defined as those 60 years and older or persons with comorbidities, and (2) all eligible five years and older. For these groups, either no new variant emerged thus the same infectiousness and severity as the Omicron variant was assumed (solid curves) or 25% more infectious and immune-evading novel variants, with Omicron-level variant severity, emerged annually (dashed curves) with horizontal dashed lines indicating initial variant emergence. Shaded areas represent stochastic uncertainty surrounding projections. Cumulative number and timing of vaccine doses are shown for each scenario. Seasonality is illustrated in the bottom row where red shading indicates warmer spring and summer seasons, blue cooler fall and winter seasons, and white the seasonal transition. See Supplementary Information Figures S3A−S5B.

Similar trends were observed when variants that are 25% more infectious and immune evading, but similar in severity as the Omicron variant, emerged annually and became dominant, although overall reductions in hospitalisations were more modest than when no variant emerged. Over the two-year study period, if variants were to emerge biannually then 15% (95% CI 3−25%) of hospitalisations could be averted if only those most vulnerable were boosted yearly, and 44% (95% CI 36−64%) if those eligible were boosted yearly compared with no boosters. Increasing booster frequency to every six months could avert 35% (95% CI 28−43%) or 40% (95% CI 31−45%) of COVID-19-related hospitalisations if those most vulnerable or those eligible were boosted, respectively. Importantly, boosting 98% of those eligible (those five years and older who received vaccine doses one and two) actually protects those most vulnerable 60 years and older or those with comorbidities more than only boosting this vulnerable group as shown in Figure 6.

We further explored the effect of a viral variant emerging conservatively every six months, assuming each new variant is 25% more immune-evading and 25% more infectious than the previous dominant variant, but with the same level of severity as the Omicron variant (Supplementary Information Figures S10A and S10B show prevalence for each variant over time). Under these conditions, we also estimate a substantial benefit of annual boosters for all eligible people followed by diminishing returns of boosting more frequently than once per year. However, the relative differences between these strategies are much less pronounced, as the new variants partially evade any previously acquired immunity after both natural infection and vaccination, leading to more overall SARS-CoV-2 infections. We show the cumulative projected impact on SARS-CoV-2 infections and COVID-19-related hospital admissions with no emerging variant or new variants emerging every twelve or six months (Figure 2). This reinforces that regardless of the presence or frequency of new emerging variants, while there is benefit to boosting those most vulnerable annually ahead of the cooler seasons, administering annual boosters to all those eligible shows the biggest gain in infections and hospital admissions averted (Figure 2, blue curves). Whereas boosting every six months is predicted to have the biggest benefit, based on the additional vaccine booster doses needed to boost twice as often (every six months), we estimated diminishing returns (Figure 2, green and purple curves).

**Figure 2.**
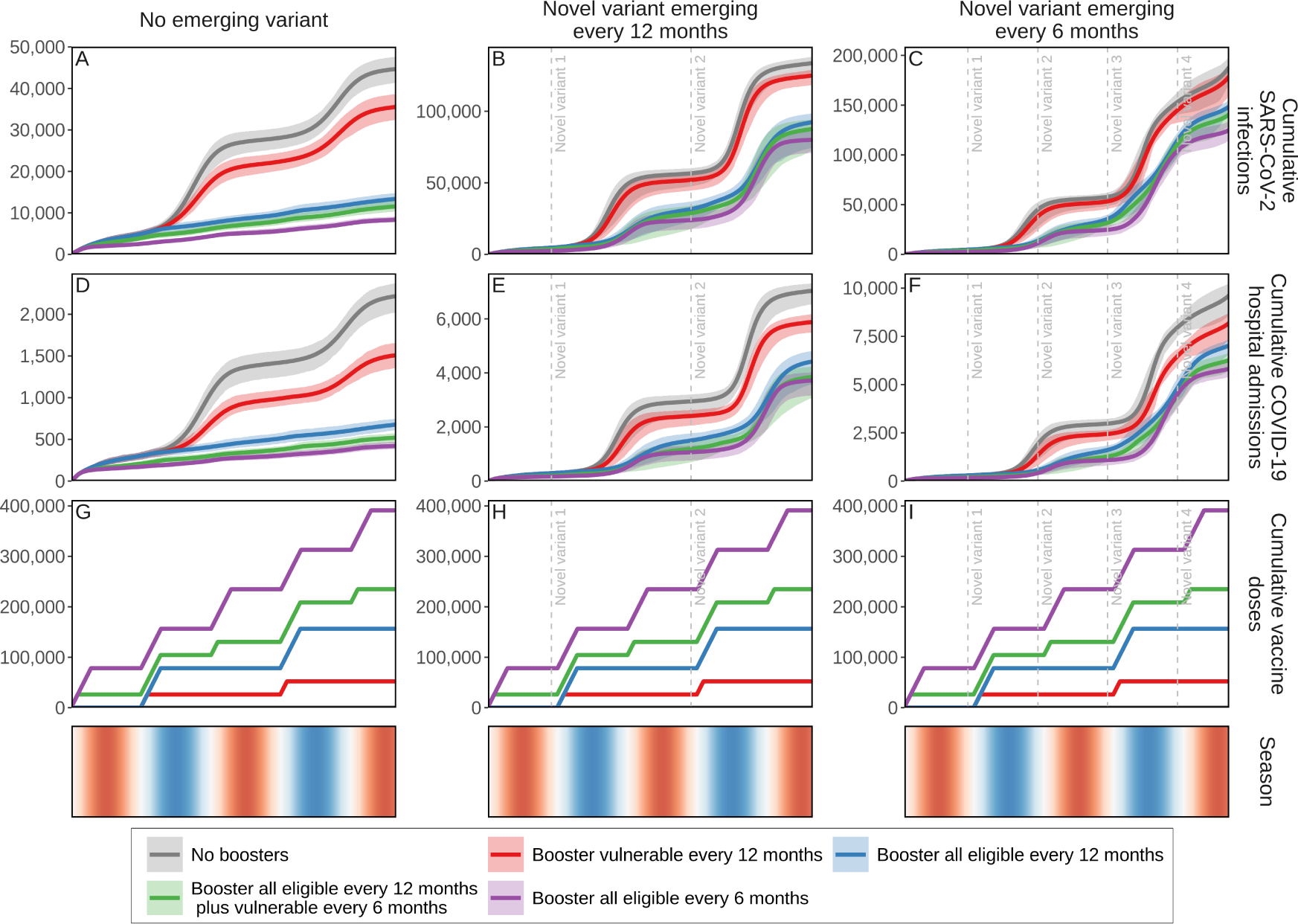
Cumulative projected impact on SARS-CoV-2 infections and COVID-19-related hospital admissions for just over a two-year period in a simulated population of 100,000 individuals with those who previously received vaccine doses one and two being eligible to receive a first-generation COVID-19 vaccine booster dose. Individuals who received boosters every twelve or six months are aggregated in two groups(1) those most vulnerable, defined as those 60 years and older or persons with comorbidities, and (2) all eligible five years and older. For these two groups, scenarios were designed either with no new emerging SARS-CoV-2 variants, assuming the same infectiousness and severity of the Omicron variant (panels A, D, and G), or with 25% more infectious and immune-evading novel variants with the same severity as the Omicron variant emerging annually (panels B, E, and H) or biannually (panels C, F, and I) with initial emergence indicated by horizontal dashed lines. Shaded areas represent the stochastic uncertainty surrounding projections. Cumulative number and timing of vaccine doses are shown for each scenario. Seasonality is illustrated in the bottom row where red shading indicates warmer spring and summer seasons, blue cooler fall and winter seasons, and white seasonal transitions. See Supplementary Information Figures S6, S7, and S10−S14B.

We also examined whether lower booster uptake of 85% coverage among those 50 years and older or persons with comorbidities and 50% among those 5−49 years of age, will have a substantial impact on the public health burden over a two-year period and found that when no new variants emerge, COVID-19-related hospitalisations could be reduced by 60% (95% CI 53−65%) (Figure 3 panel C dark blue curve) at this lower booster coverage compared with 70% (95% CI 65−73%) when 98% of those eligible (those eligible include 93% of those 60+ or with comorbidities, 90% of 50−59-year-olds, 80% of 30−49-year-olds, and 75% of 5−29-year-olds who received doses one and two) are boosted annually (Figure 3 panel C light blue curve). Assuming a new variant emerges annually, we find that hospitalisations could be reduced by 37% (95% CI 30−48%) with universal boosting (98% coverage of those eligible), dropping to 26% (95% CI 18−32%) with lower coverage (Figure 3 panel D dark and light blue curves, respectively).

**Figure 3:**
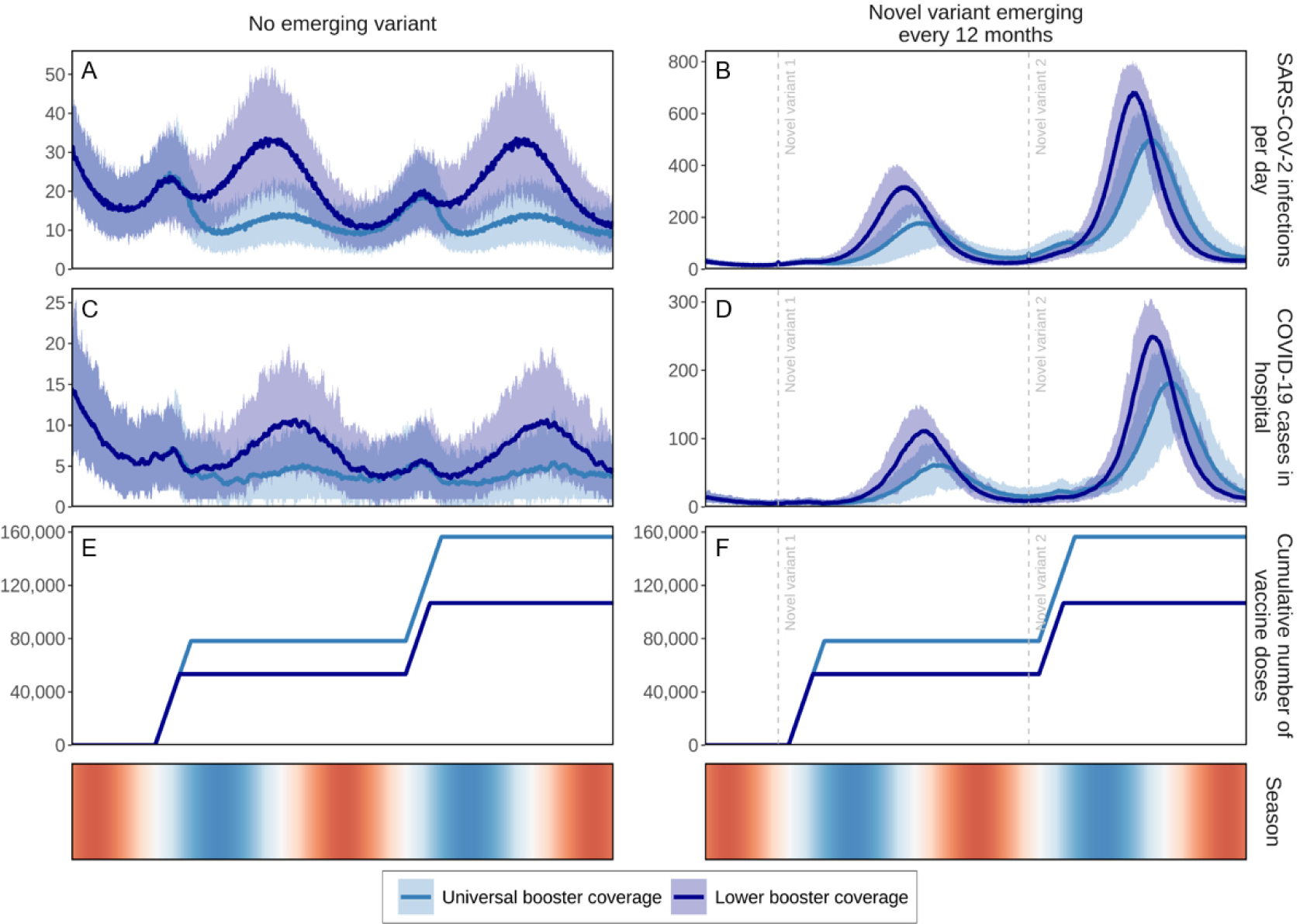
Daily projected impact on SARS-CoV-2 infections and COVID-19-related hospital admissions over two-years in a simulated population of 100,000 individuals with first-generation COVID-19 vaccine boosters administered annually. Boosters were administered at universal coverage (98% of those eligible to receive a booster dose (including 93% of those 60 years and older or persons with comorbidities, 90% of 50−59-year-olds, 80% of 30−49-year-olds, and 75% of 5−29-year-olds who received doses one and two) and lower coverage levels (85% of those eligible who are 50 years and older or who are living with comorbidities, and 50% of those eligible 5−49 years of age). For these two groups, either no new SARS-CoV-2 variants emerged, assuming the same infectiousness, immune-evading capacity, and severity of the Omicron variant (panels A, C, and E) or novel variants with 25% more infectiousness and immune-evading capacity than the previously dominant variant but with the same level of severity as the Omicron variant, emerged every twelve months, with initial variant emergence indicated by horizontal dashed lines (panels B, D, and F). Shaded areas represent stochastic uncertainty surrounding projections. Cumulative number and timing of boosters are shown for each scenario. Seasonality is illustrated in the bottom row where red shading indicates warmer spring and summer seasons, blue cooler fall and winter seasons, and white the seasonal transition. See Supplementary Information Figure S7.

To minimise SARS-COV-2 infections and COVID-19-related hospital admissions when a new variant of concern emerges annually ahead of the winter season, we found that the best time to deliver annual COVID-19 booster doses is three to four months before peak winter when people tend to remain indoors most often and their number of contacts increases as does their probability of SARS-CoV-2 transmission (Figure 4 and Supplementary Information Figure S16A). The seasonality profile as illustrated in Supplementary Information Figure S2, including the seasonality forcing scaler varied in the sensitivity analysis shown in Figure 5, will affect these results. In addition, for this modelling study by default it was assumed booster vaccines were rolled out to 1.5% of the population every day. No effect was observed if the daily boosting capacity was reduced to 0.75% or increased to 2.25% (results not shown in Figure 5).

**Figure 4:**
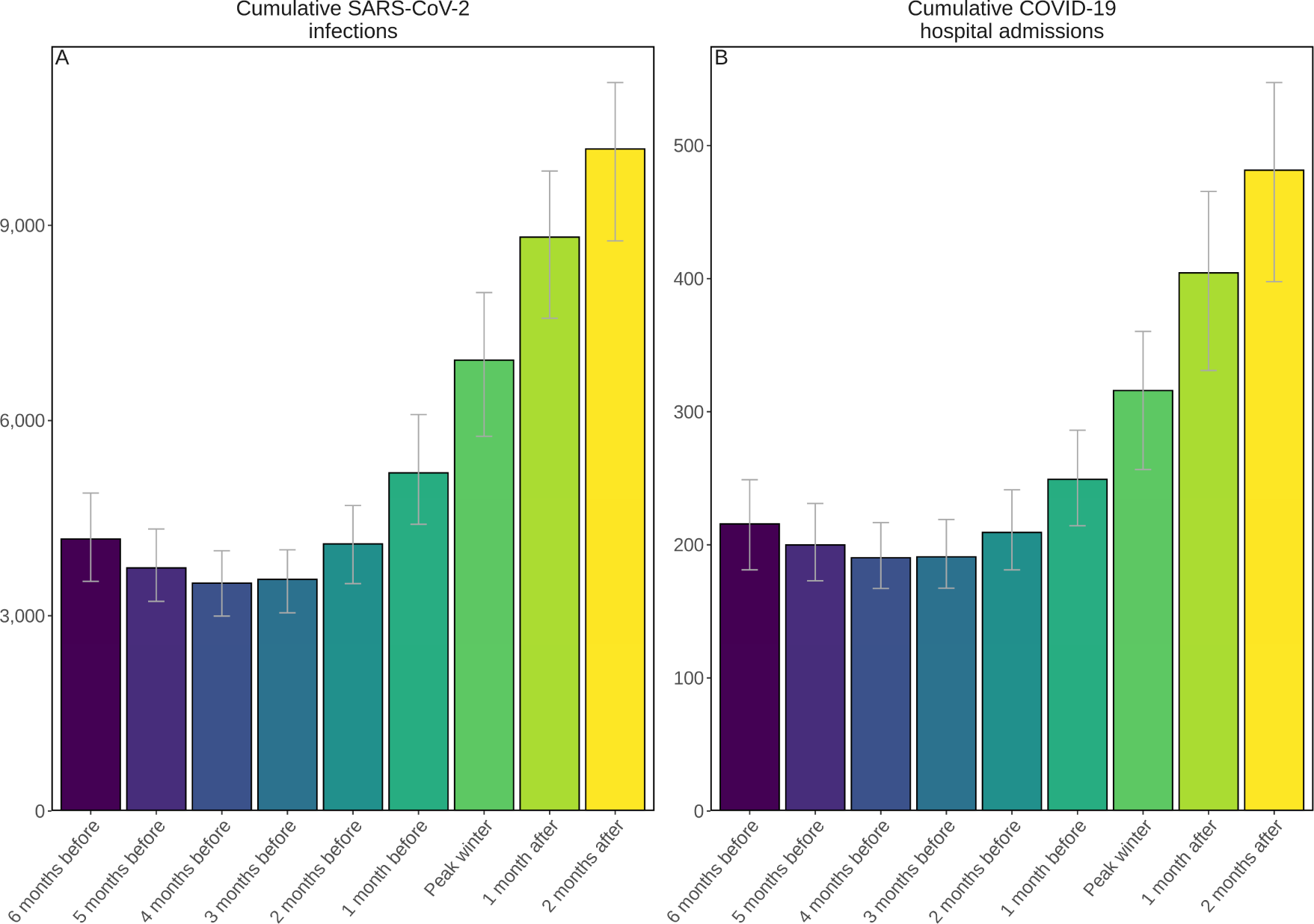
Projected cumulative annual impact on new SARS-CoV-2 infections (panel A) and COVID-19-related hospital admissions (panel B) of delivering vaccine boosters to all those eligible five years of age and older from either six months before to two months after peak winter temperatures assuming a 25% more infectious and immune-evading novel variant but with the same severity as the Omicron variant emerges prior to the fall season. Error bars represent the uncertainty in model projections. See Supplementary Information Figures S8, S16A, and S16B.

**Figure 5:**
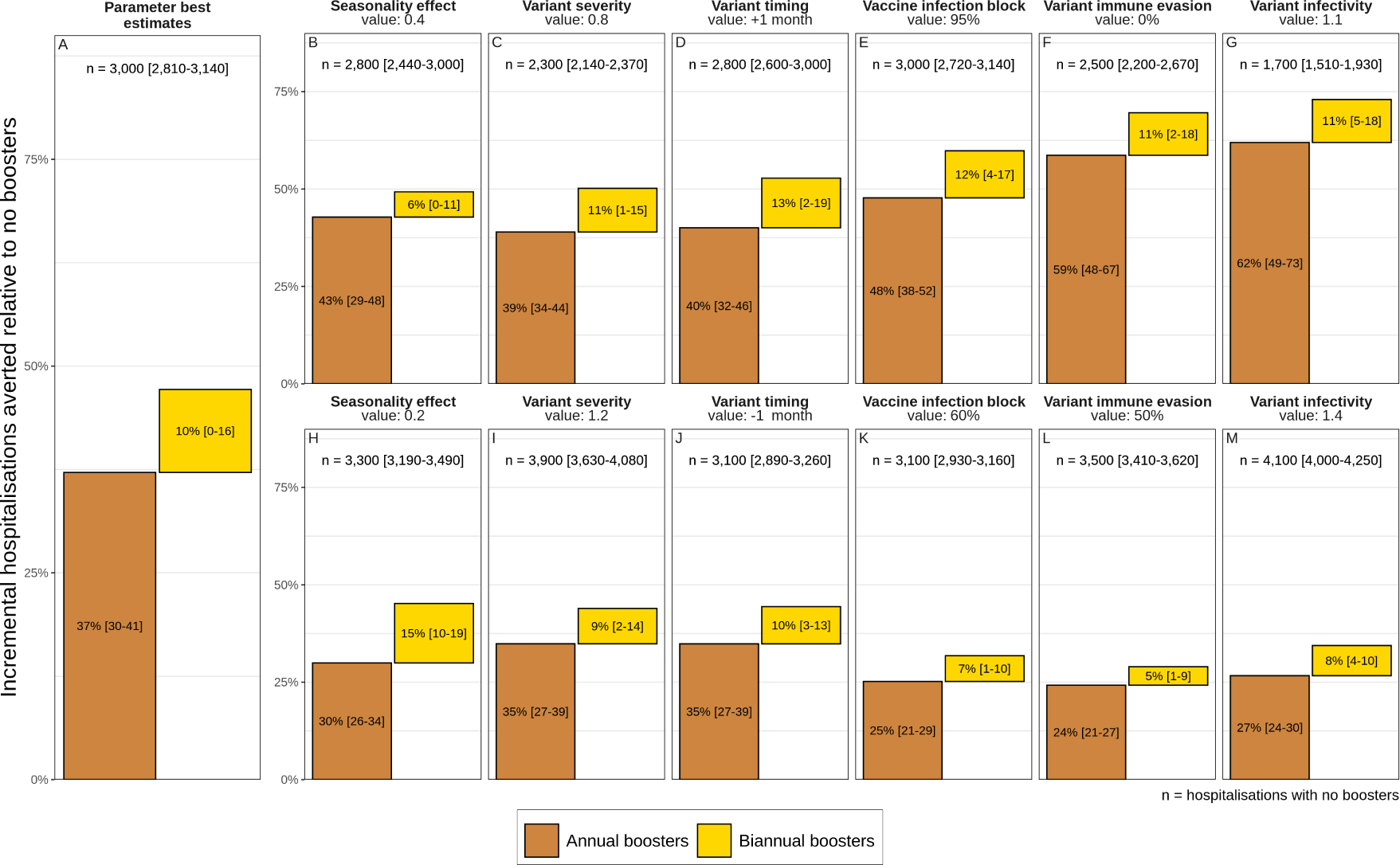
Projected percentage incremental hospitalisations averted over a two-year period by annual (brown bars) and biannual (yellow bars) COVID-19 vaccine boosters for those eligible under varying seasonality, emerging variant severity, variant timing, vaccine infection-blocking efficacy, emerging variant immune-evading capacity, and emerging variant infectivity from best estimate values as listed in Table 1. See Supplementary Information Figures S15 and S17.

**Figure 6:**
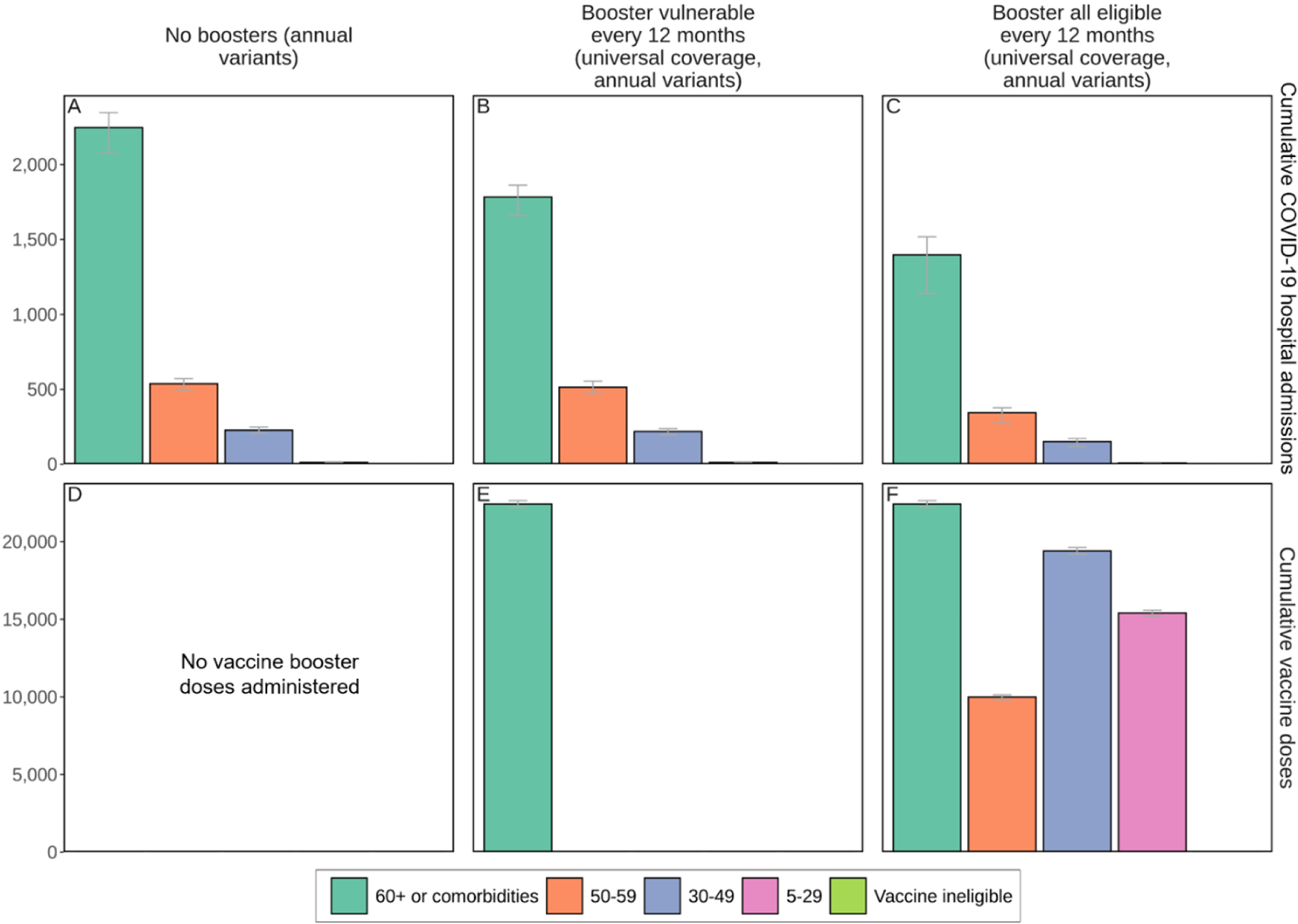
Cumulative projected impact of administering COVID-19 vaccine boosters to those most vulnerable or to all those eligible every twelve months on SARS-CoV-2 infections and COVID-19- related hospital admissions by age and risk group over a two-year period in a simulated population of 100,000 individuals when new SARS-CoV-2 variants with 25% more infectiousness than previously dominant variants emerged annually. Cumulative numbers of vaccine doses are shown in panels D-F. Error bars represent the stochastic uncertainty in model projections. See Supplementary Information Figure S9.

For all key parameters that were varied, annual boosting consistently provides the majority of reductions of COVID-19-related hospital admissions over the two-year study period. Higher variant infectivity and higher immune-evading capacity most reduced the effect of boosters (Figure 5 panels L and M). The largest relative effect of biannual boosting was observed under lower seasonal effect (Figure 5 panel H), in which case biannual boosting is responsible for half the percentage of hospitalisations averted (15% (95% CI 10−19%)) compared with annual boosters (30% (95% CI 26−34%)).

As an indication of cost-effectiveness, we found that more hospitalisations could be averted per booster dose through annual boosting of those most vulnerable versus all those eligible over the study period (Supplementary Information Figure S17). This finding was robust even when key model parameters were varied, again showing diminishing returns per dose when boosting frequency is increased to every six months. However, only boosting those most vulnerable may not be sufficient to prevent exceeding health system capacity thresholds.

## Discussion

To minimise COVID-19-related hospitalisation admissions over the next two years, our model-based evidence showed that vaccine boosters should be provided for all those eligible (those previously receiving two doses) at least three to four months ahead of the winter season whether or not new variants of concern emerge and become dominant. Our findings are consistent with those reported for a study conducted in the UK, which also recommended that antiviral supply should be ready to prioritise early treatment of those most vulnerable as soon as they test positive for SARS-CoV-2 infection as vaccines may be less effective in vulnerable populations.^14^ We found that, under our current assumptions of natural and vaccine induced waning, only boosting those most vulnerable (defined as those 60 years of age or older or people living with comorbidities) may be insufficient to ensure reduced stress on health systems.

Whereas increasing the frequency of boosters to every six months (to those most vulnerable, or all those eligible) showed diminishing returns. Importantly, we showed that boosting all those eligible protects those most vulnerable more than only boosting this vulnerable group, with important vaccine strategies implications for the forthcoming Northern 2022 winter. This strategy will also be crucial to consider noting the early and large influenza epidemic ongoing during the Australian 2022 winter, which has increased health system stress in the country. Therefore, efforts should be made to reduce COVID - related stress on health systems ahead of the anticipated influenza season in the Northern hemisphere. These results were robust when key model parameters were varied. However, for all results, the more frequently variants with mounting infectiousness emerge, the less the effect vaccine boosters will have, regardless of whether boosters are administered annually or biannually.

Resources needed to administer booster doses more often and more extensively should be measured against the predicted benefit to public health and health systems capacity. These include dose supply (including global equity in distribution), effort, and cost, required to increase the frequency. Should the number of COVID-19-related hospital admissions averted per booster dose administered be used as a proxy metric for cost-effectiveness, then more hospitalisations could be averted per booster dose through annual boosting of those most vulnerable versus all eligible, an indication of cost-effectiveness. However, only boosting those most vulnerable may be insufficient to prevent health system risk.

Vaccination hesitancy and fatigue must also factor into this assessment and be considered as part of vaccine booster campaigns and messaging and lessons can be learned from seasonal influenza strategies. Even if there were not challenges surrounding vaccine hesitancy and repeated booster fatigue, we cannot boost indefinitely against SARS-CoV-2 infection and COVID-19 disease, and regardless, evidence from this study suggests that more frequent boosting to every six months has diminishing returns on investment (Supplementary Information Figure S17). Should booster doses not be available or not be accepted particularly as more infectious as potentially more severe subvariants emerge, hospital admissions may once again overwhelm the health system. This compounded with the potential for double epidemics with influenza and respiratory syncytial virus (as has been reported in Australia^15^), will escalate the need for stronger preparedness for such eventualities given the high mutation rate of SARS-CoV-2.^16^ Therefore it is prudent for countries to continue targeting and supporting vaccination boosting campaigns for all. Since it is anticipated that more infectious and more immune escaping variants will continue to emerge, existing vaccines may become insensitive to new variants, which reinforces the need for effective adapted and possibly multivalent vaccines with longer durations, coupled with plans for adjusted and improved strategies over the years.

While it is difficult to project cost-effectiveness, the impact per booster dose can be used as a proxy for health savings. We find that more hospitalisations could be averted over a two-year period per booster dose through annual boosting of those most vulnerable than all those eligible even when assumption values for variant infectivity, immune-evading capacity, vaccine infection blocking, seasonality effect, variant severity, and variant timing are increased or decreased within assumed bounds, and under assumptions for waning immunity and study settings.

### Vaccine targets and inequity of vaccine distribution

While the WHO target to fully vaccinate (with two doses) 40% of the world’s population against SARS-CoV-2 by the end of 2021 was missed, coverage reached 60% by the end of May, leaving only a month remaining to achieve the 70% mid-2022 coverage target.^17^ While it is important to consider optimal booster strategies, the WHO warns that broad-based COVID-19 booster programmes, including delivering additional booster vaccinations to groups at lower risk of developing severe disease may prolong the global pandemic by diverting supply to nations with already high coverage and therefore away from nations with low coverage, giving the virus more chance to spread and mutate.^10^ Given that 2.7 billion people had still not even received their first dose of the COVID-19 vaccine as of the end of May 2022,^18^ WHO experts have urged prioritising first doses over boosters.^19^ Similarly, Gavi, the Vaccine Alliance, advocates for the global community to focus on ensuring every adult receives at least one dose in 2022.^18^

### Preparedness for future waves

While the number of confirmed COVID-19-related deaths may be underreported,^20^ the healthcare system now has more collective experience treating those with severe disease than it did during previous waves of COVID-19. Medical oxygen supplies and distribution have been strengthened in many countries.^15^ In addition, oral antivirals to treat COVID-19 have been authorised for inpatient and home use in several countries, as well hospital administered injectable monoclonal antibodies are now available. These pharmaceutical interventions, together with mask usage and physical distancing measures being observed to varying degrees across settings, and coupled with increasing vaccination rates and natural immunity from previous infection may make certain settings better positioned to mitigate and respond to the subsequent COVID-19 waves. Nevertheless, with continuing spread of Omicron subvariants, even if Omicron infection causes less severe disease, should cases spike again, a smaller proportion of a large number of cases needing hospitalisation may again lead to health system stress.

While increasing vaccination coverage is a good solution to reducing the burden from COVID-19, a sizable proportion of the population continues to refuse vaccination in high-, middle-, and even low-income settings.^21^ Even though testing rates have plateaued,^20^ meaning cases are likely being underreported, ongoing SARS-CoV-2 transmission will inevitably result in the emergence of new SARS-CoV-2 variants or subvariants of concern. This is why it is important to be strategic with COVID-19 vaccination campaigns. As we cannot continue to boost for COVID-19 indefinitely, in efforts to avoid vaccination fatigue and to better protect against new variants, groups are working to develop second-generation vaccines that are multivalent, meaning they protect better against more than one SARS-CoV-2 variant, and will remain more effective, longer. Clinical trial results for a new Omicron-specific bivalent booster candidate (mRNA-1273.214) are expected later this year.^22^ Under the booster strategy recommended in this study, more effective (as shown in the sensitivity analysis conducted in this study) and longer lasting second-generation booster vaccines will avert even more infections and hospital admissions than were predicted using first-generation boosters.

### Study limitations

As with all simulation studies our results are based on several assumptions and includes some limitations. Within our individual-based transmission model, transition between disease states and chance to be infected includes probabilities for infection, transmission, and disease severity, based on studies cited in ^6^. These probabilities will influence model outcomes and our conclusions on vaccination so sensitivity analysis was conducted on several parameters. First, we identified that Omicron’s potential severity had little effect on the number of new cases projected, also because no impact on viral load was assumed. Higher viral load in the model is associated with increased transmissibility. Second, values higher or lower than 1.0 for the effective reproduction number at the start of the winter period, prior to introduction of Omicron, may influence peak hospital admissions estimates; however, the relative impact of vaccination is not sensitive to this parameter. Third, this study only considers direct admissions to hospital due to COVID-19, but does not consider COVID-19-related admissions due to certain long-COVID health consequences. Therefore, there is the potential for additional health system burden not modelled here. Fourth, we did not model the effect of any NPIs, but we assume re-implementing NPIs could be considered if thresholds for case numbers or health system capacity were reached. We also acknowledge that booster vaccines began to be rolled-out as early as fall 2021 in certain countries, whereas we simulated booster administration starting in spring 2022. Last, depending on emerging variant profile and vaccine coverage levels, should naturally acquired- and/or vaccine-induced-immunity have different protection levels and/or should immunity wane differently (speed, decay type) than what was assumed, then the population would be more or less protected and our projected outcomes may prove to be too pessimistic or too optimistic. We did not model the effect of hybrid immunity, which occurs when an individual experiences both naturally acquired and vaccine-induced immunity. Recent studies have identified hybrid immunity to provide a higher level of protection against infection and severe disease than we assumed in the model.^23–25^ Since new SARS-CoV-2 variants of concern, and therefore infections, will continue to emerge, and vaccinations will continue to need to be implemented, the prevalence of hybrid immunity will increase. This will affect population susceptibility, especially in the long-term. As more is known about hybrid immunity to SARS-CoV-2 infection and COVID-19 disease, this should be considered as part of future modelling studies, and when interpreting these results.

To minimise the public health burden of COVID-19 and the associate and health system risk from, vaccine boosting strategies and policy decisions must account for rollout constraints and cost-effectiveness. This study provides timely needed quantitative evidence for decisions around future boosting strategies.

## Data Availability

No datasets were generated or analysed during this study. Data informing model parameters are described herein and in Shattock et al.6 Open-access model and figure code are available at https://github.com/SwissTPH/OpenCOVID. Data underpinning figures are available upon request.

## Ethics approval and consent to participate

Not applicable.

## Consent for publication

All authors gave final approval for publication. No other consent is required.

## Availability of data and materials

No datasets were generated or analysed during this study. Data informing model parameters are described herein and in Shattock et al.^6^ Open-access model and figure code are available at https://github.com/SwissTPH/OpenCOVID. Data underpinning figures are available upon request.

## Authors’ contributions

SLK, MAP and AJS conceived the study with input from EALR, and MR. AJS further developed the model with input from SLK, EALR, MR, and MAP. AJS and SLK performed the analyses, prepared the figures, and conducted model and analysis validation. SLK drafted the manuscript together with AJS. All authors contributed to interpreting results and editing the manuscript.

## Acknowledgements

The authors want to acknowledge and thank the members of the Disease Modelling Unit, Swiss Tropical and Public Health Institute, specifically Dr Nakul Chitnis for his input. Simulations were performed at sciCORE (http://scicore.unibas.ch/) the scientific computing facility at the University of Basel.

## 1. Supplementary methods

### 1.1. Disease prognosis and effect of variant properties

Our individual-based model, OpenCOVID, is a dynamic stochastic discrete-time model of SARS-CoV-2 transmission and COVID-19 disease progression and response.^1^ The model simulates viral transmission between infectious and susceptible individuals that come in contact through an age-structured, small-world network. The probability of transmission in each exposure stage is influenced by the infectiousness of the infected individual, the immunity of the susceptible individual (acquired through previous infection and/or through vaccination), and a background seasonality pattern (reflecting a larger proportion of contacts being in closer contact indoors as the temperatures became cooler). Infectiousness is a function of variant infectivity and time since infection. Once infected, a latency period is followed by a pre-symptomatic stage, after which an individual can experience asymptomatic, mild, or severe disease. Severe cases can lead to hospitalisation, intensive care unit admission, and ultimately death (see Supplementary Table S1 for further details on prognosis probabilities by age, following infection with SARS-CoV-2 Omicron variant). Recovery after infection leads to development of immunity. This immunity is assumed to wane over time as illustrated in Figure S1, with the risk of new infection depending on the probability of exposure and properties of existing and potential novel SARS-COV-2 variants (i.e., infectiousness and immune evading profile). The model has the capacity to represent a number of containment measures, including non-pharmaceutical interventions such as physical distancing and facemask usage, testing strategies such as test-diagnose, isolate or quarantine, mass testing, and contact tracing, and pharmaceutical interventions, such as vaccination and treatment. A detailed description of the OpenCOVID model is provided in ^1, 2^. Open access source code for the OpenCOVID model is publicly available at https://github.com/SwissTPH/OpenCOVID.

### 1.2. Vaccine-induced and naturally acquired SARS-CoV-2 immunity profiles

**Figure S1:**
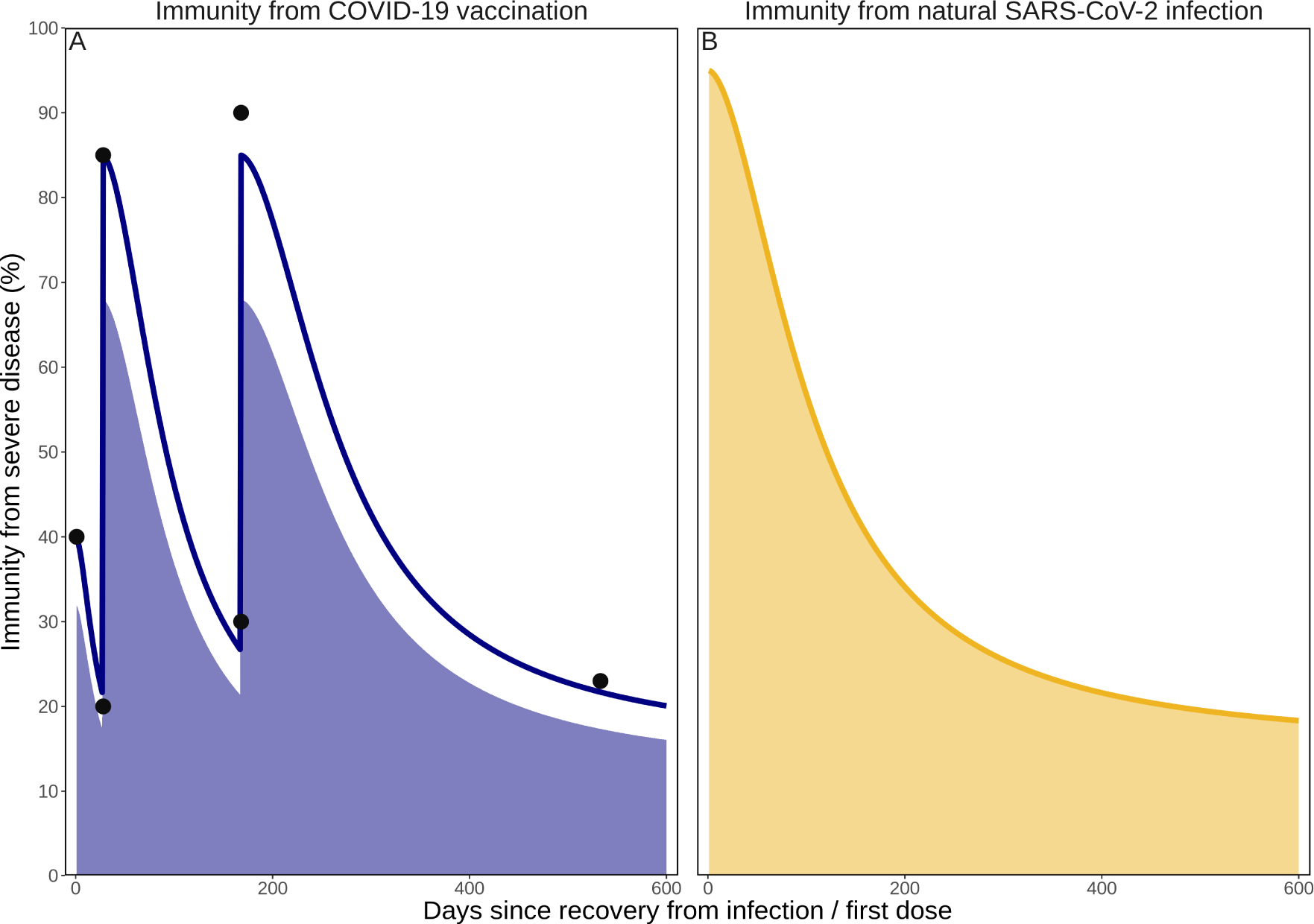
Profiles of COVID-19 vaccine induced immunity (panel A) and naturally acquired immunity following infection with SARS-CoV-2 (panel B) as described in the *Vaccination strategy* section of the *Methods* in the main text. Points indicate initial vaccine efficacy with exponential decay, with subsequent booster dose efficiency rebounding with an identical decay (based on ^3^).

### 1.3. Disease prognosis and effect of variant properties

The probabilities of 1) a symptomatic case developing severe disease, 2) a severe case becoming critical, and 3) a critical case ultimately leading to death are defined as functions of age (Table S1). Additionally, individuals with severe comorbidities also experience an increased probability in developing severe disease. In our simulations, about 33% of the total population is considered most vulnerable, i.e. at highest risk of developing severe disease, which includes those 60 years and older or those with one or more severe comorbidities. In this study we used continuous functions fitted to the discrete probabilities reported in ^4^, updated to represent the additional risk of severe disease from infection with the SARS-CoV-2 Omicron variant (B.1.1.529)^5–7^. Table S2 shows the multiplicative factors used to represent Omicron, relative to wild type SARS-CoV-2 for which the prognosis probabilities in ^4^ were estimated. In addition to age-related risk, the probability that an infected individual will develop severe symptoms can also be scaled by the severity factor corresponding to the viral variant to which they were exposed.

For vaccinated individuals that become infected (noting that the infection-blocking action of the vaccine reduces the probability of infection), the probability of developing severe disease is reduced by the disease-blocking action of the vaccine. The level to which the probability of severe disease is reduced is dependent upon the level of immunity at the time of infection. Vaccine-induced immunity is assumed to wane over time (Figure S1) and can be further decreased if exposed to a variant with immune-evading capacity.

**Table S1:** Prognosis probabilities by age, following infection with SARS-CoV-2 Omicron variant

| Age group (years) | Asymptomatic | Mild disease | Severe disease | Critical disease | Death |
| --- | --- | --- | --- | --- | --- |
| 0->10 | 33.00% | 67.00% | <0.01% | <0.01% | <0.01% |
| 10->20 | 33.00% | 66.98% | 0.02% | <0.01% | <0.01% |
| 20->30 | 32.97% | 66.81% | 0.21% | 0.01% | <0.01% |
| 30->40 | 32.94% | 66.52% | 0.51% | 0.03% | <0.01% |
| 40->50 | 32.65% | 64.35% | 2.77% | 0.24% | <0.01% |
| 50->60 | 31.73% | 58.00% | 8.91% | 1.36% | 0.01% |
| 60->70 | 29.54% | 45.44% | 18.89% | 5.99% | 0.14% |
| 70->80 | 26.74% | 33.23% | 22.64% | 15.21% | 2.17% |
| 80-90+ | 24.22% | 24.94% | 15.66% | 21.55% | 13.63% |

**Table S2:** Infectivity and severity of SARS-CoV-2 variants of concern (VOC) relative to the previously dominant variant

| VOC | Lineage name | Date of designation<sup>8</sup> | Variant overtaken | Infectivity | Severity |
| --- | --- | --- | --- | --- | --- |
| Wild-type |  | 31 Dec 2019 |  |  |  |
| D614 | D614 | 15 Mar 2020 | Wild-type | 1.00 | 1.00 |
| G614 | G614 |  | D614 | 3.00<sup>2</sup> | 1.00 |
| Alpha | B.1.1.7 | 18 Dec 2020 | G614 | 1.64 (95% CI 1.32–2.04)<sup>4</sup> | 1.67 (95% CI 1.34–2.09)<sup>4</sup> |
| Delta | B.1.617.2 | 11 May 2021 | Alpha | 1.50<sup>5</sup>–6.00<sup>6</sup> | Hospitalisation: 1.85 (95% CI 1.39–2.47)<sup>7</sup><br>ICU admission: 2.34 (95% CI 1.64–3.31)<sup>9</sup><br>Death: 1.32 (95% CI 0.47–2.30)<sup>9</sup> |
| Omicron | B.1.1.529 | 26 Nov 2021 | Delta | 3.20 (95% CI 2.00–5.00)<sup>10</sup> | Hospitalisation among 60–69-year-olds: 0.75<sup>11</sup> |

### 1.4. Seasonality

Seasonality is assumed to follow a scaled cosine function, oscillating between a minimum point (-1), representing minimum seasonal forcing during the peak summer period, and maximum seasonal forcing during the peak winter period (1). The best estimate for the seasonal scaling factor, 0.3, was taken from a previous study assessing the COVID-19 epidemic in Switzerland.^1^ This previous study simultaneously fit this seasonal scaling factor parameter along with several other parameters to align the OpenCOVID model to epidemiological data. To assess the effect of the seasonal scaling factor on model outcomes, two additional values for the peak summer effect were modelled in a sensitivity analysis (see Figure 5 from the main text): 0.2 (yellow curve, representing a less seasonal, more perennial setting) and 0.4 (green curve, representing a more seasonal setting).

**Figure S2:**
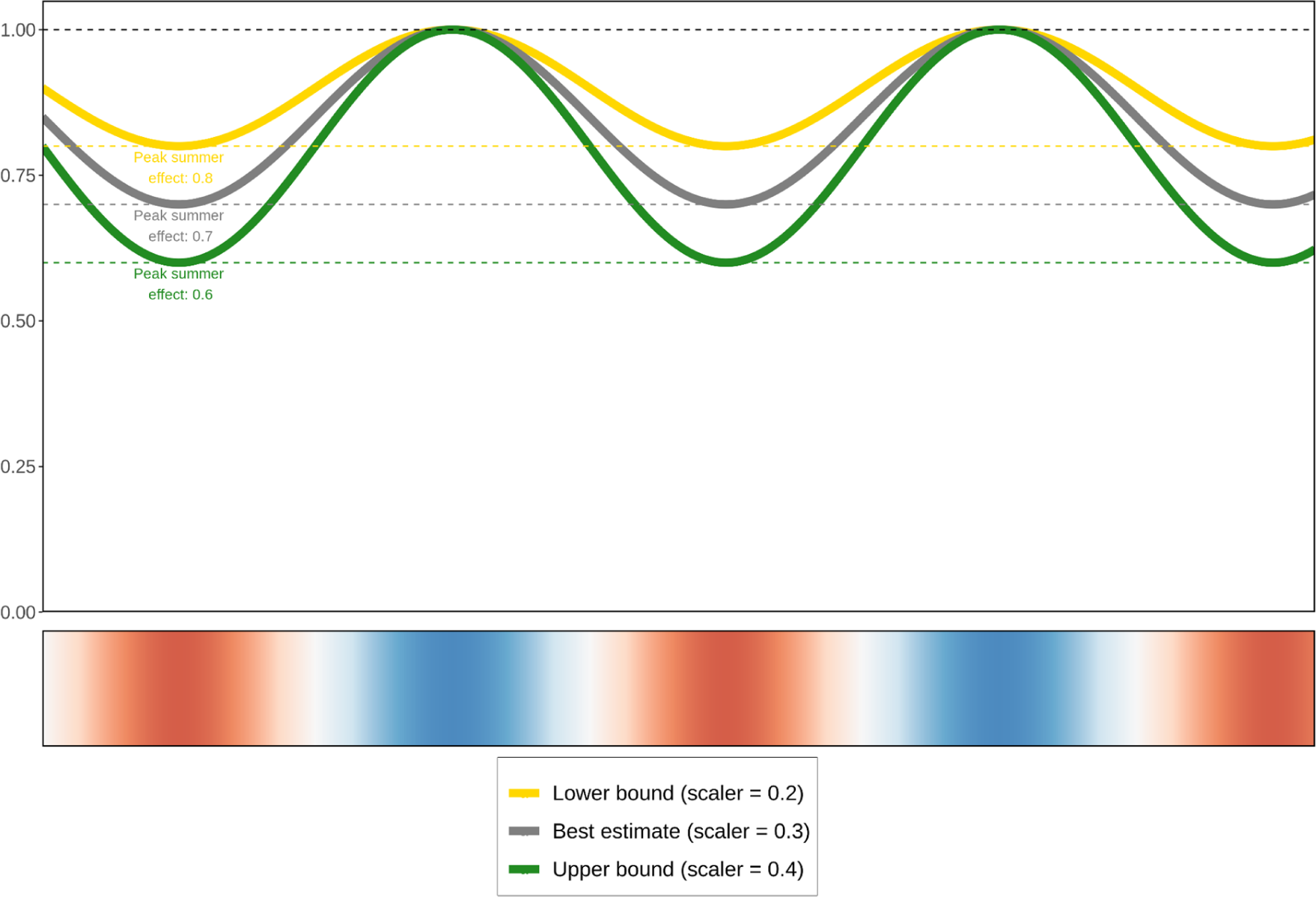
Impact of seasonal forcing scalers (best estimate (grey curve), lower bound (yellow curve), and upper bound (green curve)) on SARS-CoV-2 infectiousness per contact over a two year period. Seasonality is illustrated in the bottom row where red shading indicates the warmer spring and summer seasons, blue the cooler fall and winter seasons, and white the seasonal transition periods.

## 2. Supplementary results

**Figure S3A:**
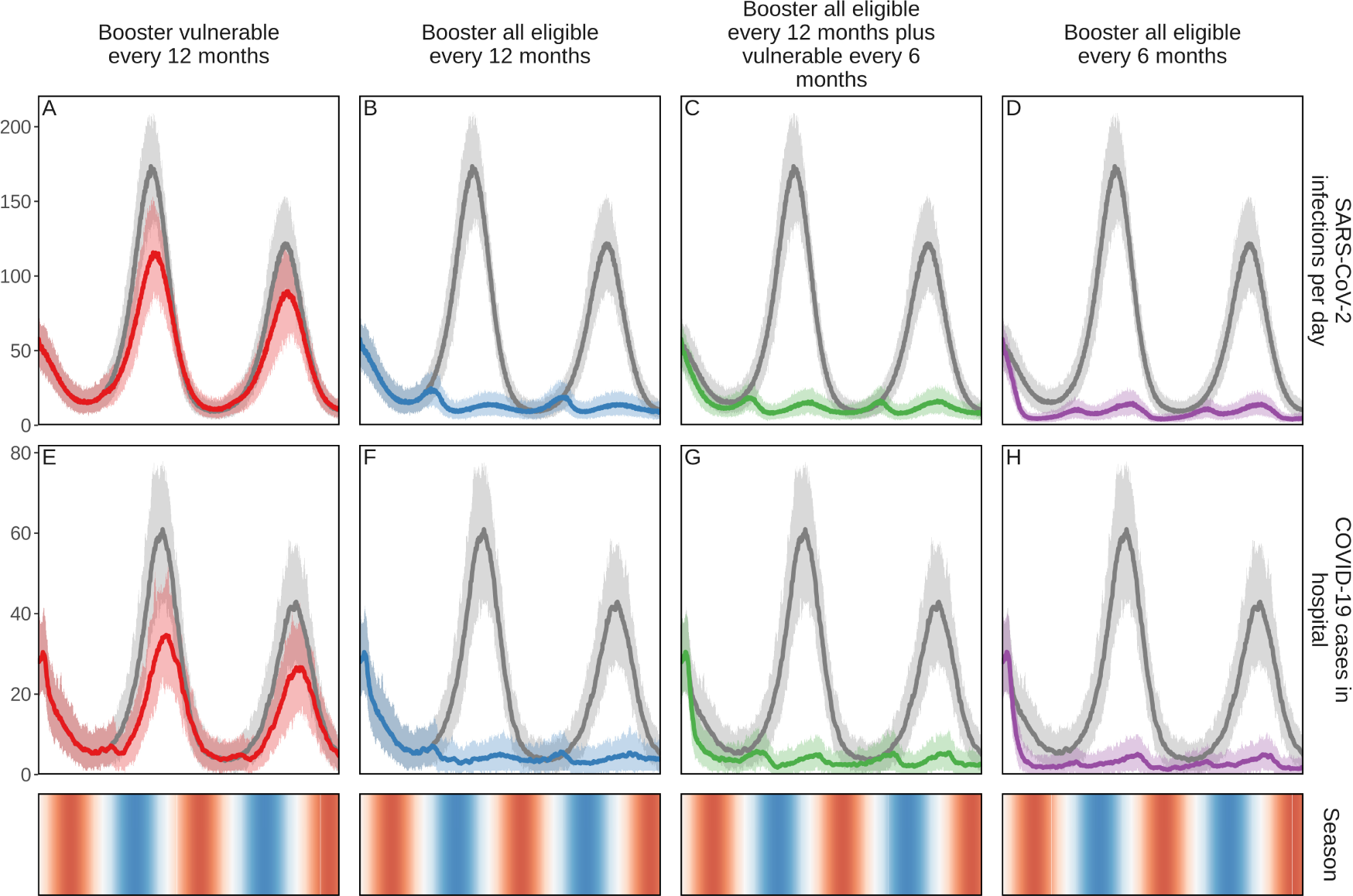
Projected impact of COVID-19 vaccine boosters on daily SARS-CoV-2 infections and COVID-19-related cases in hospital. The simulation was for just over a two-year period in a population of 100,000 individuals. Those who were modelled as having previously received vaccine doses one and two, including 93% of those 60+ or with comorbidities, 90% of 50−59 year olds, 80% of 30−49 year olds, and 75% of 5−29 year olds, are eligible to receive first-generation COVID-19 vaccine boosters. Populations who received boosters every twelve or six months are aggregated into two groups, (1) those most vulnerable (60 years and older or persons with comorbidities) and (2) all eligible (five years and older). Each vaccine scenario curve is plotted alongside the baseline scenario whereby no vaccine booster doses were administered (grey curves). In these simulations, no new SARS-CoV-2 variants emerged, and the same infectiousness and severity of the Omicron variant was assumed. Shaded areas represent the stochastic uncertainty surrounding projections. Cumulative number and timing of vaccine doses are shown for each scenario. Seasonality is illustrated in the bottom row where red shading indicates the warmer spring and summer seasons, blue the cooler fall and winter seasons, and white the seasonal transition periods. This figure is an extension of Figure 1 from the main text.

**Figure S3B:**
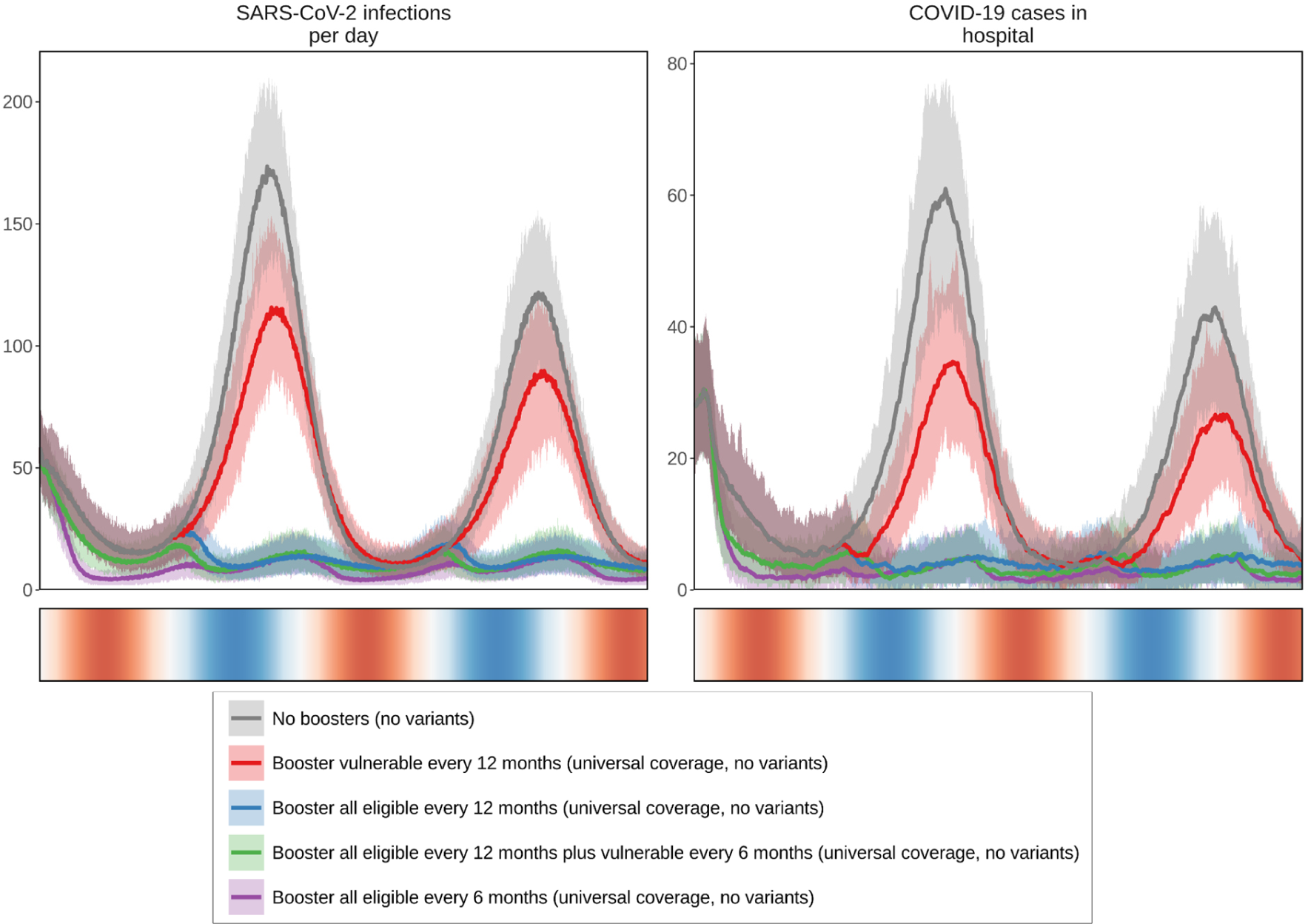
Projected daily impact on SARS-CoV-2 infections and COVID-19-related cases in hospital over a two-year period in a simulated population of 100,000 individuals. Those who were modelled as having previously received vaccine doses one and two, including 93% of those 60+ or with comorbidities, 90% of 50−59 year olds, 80% of 30−49 year olds, and 75% of 5−29 year olds, are eligible to receive first-generation COVID-19 vaccine boosters. Populations who received boosters every twelve or six months are aggregated into two groups, (1) those most vulnerable (60 years and older or persons with comorbidities) and (2) all eligible (five years and older). Vaccine scenario curves are plotted alongside a baseline scenario (grey curve) whereby no vaccine booster doses were administered (grey curves). Universal coverage of booster doses is 98% of those eligible. In these simulations, no new SARS-CoV-2 variants emerged, and the same infectiousness and severity of the Omicron variant was assumed. Shaded areas represent the stochastic uncertainty surrounding projections. Cumulative number and timing of vaccine doses are shown for each scenario. Seasonality is illustrated in the bottom row where red shading indicates the warmer spring and summer seasons, blue the cooler fall and winter seasons, and white the seasonal transition periods. This figure shows the same results as Figure S3A but with curves plotted within the same outcome panel.

**Figure S4A:**
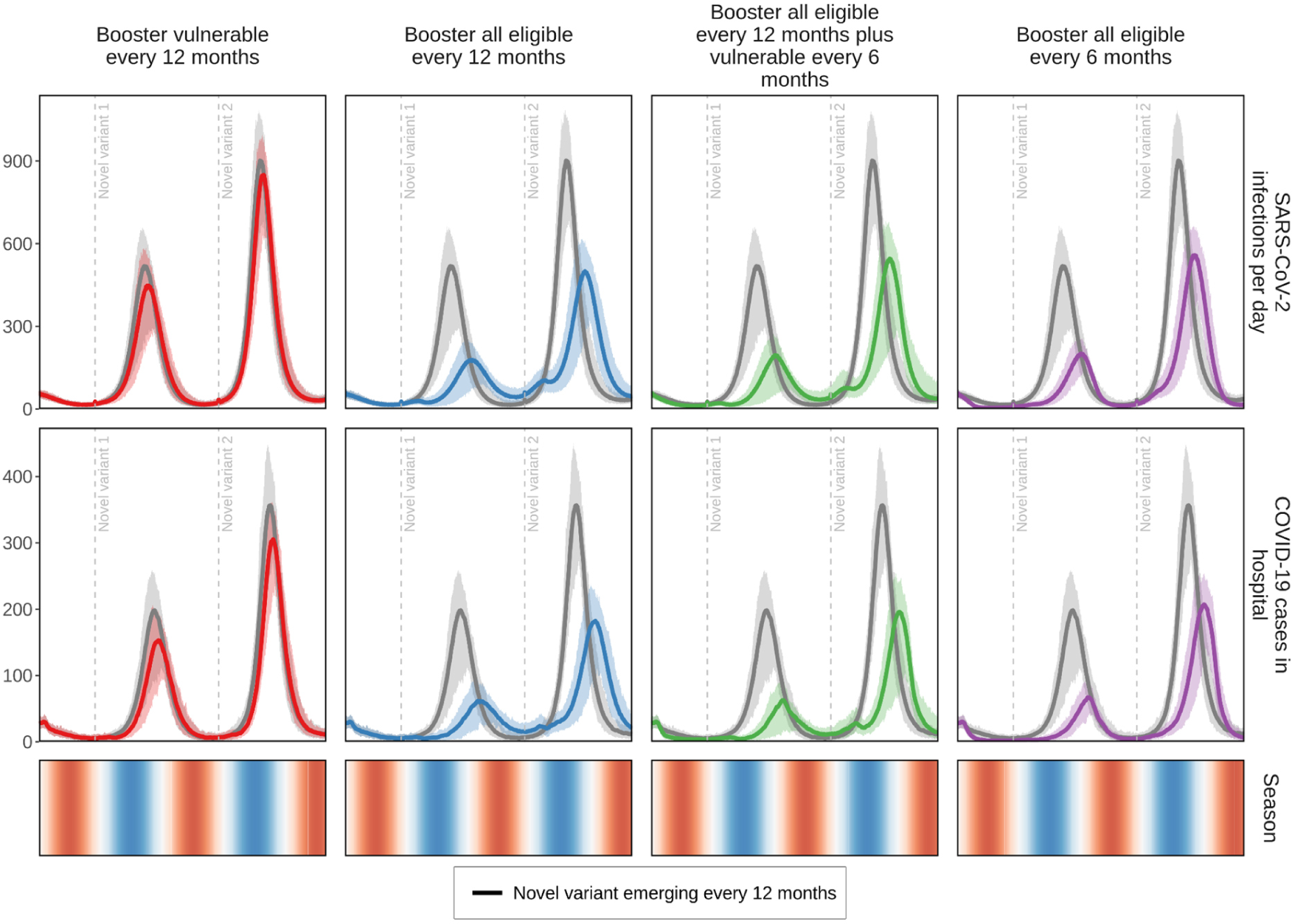
Projected daily impact on SARS-CoV-2 infections and COVID-19-related hospital admissions for just over a two-year period in a simulated population of 100,000 individuals. Those who were modelled as having previously received vaccine doses one and two, including 93% of those 60+ or with comorbidities, 90% of 50−59 year olds, 80% of 30−49 year olds, and 75% of 5−29 year olds, are eligible to receive first-generation COVID-19 vaccine boosters. Populations who received boosters every twelve or six months are aggregated into two groups, (1) those most vulnerable (60 years and older or persons with comorbidities) and (2) all eligible (five years and older). Each vaccine scenario curve is plotted alongside a baseline scenario (grey curve) whereby no vaccine booster doses were administered (grey curves). Booster dose coverage is 98% of those eligible (universal coverage). Novel SARS-CoV-2 variants with 25% more infectiousness and immune evading capacity than the previously dominant variant but with the same severity as the Omicron variant emerged annually (initial emergence indicated with horizontal dashed lines). Shaded areas represent the stochastic uncertainty surrounding projections. Cumulative number and timing of vaccine doses are shown for each scenario. Seasonality is illustrated in the bottom row where red shading indicates the warmer spring and summer seasons, blue the cooler fall and winter seasons, and white the seasonal transition periods. This figure is an extension of Figure 1 from the main text.

**Figure S4B:**
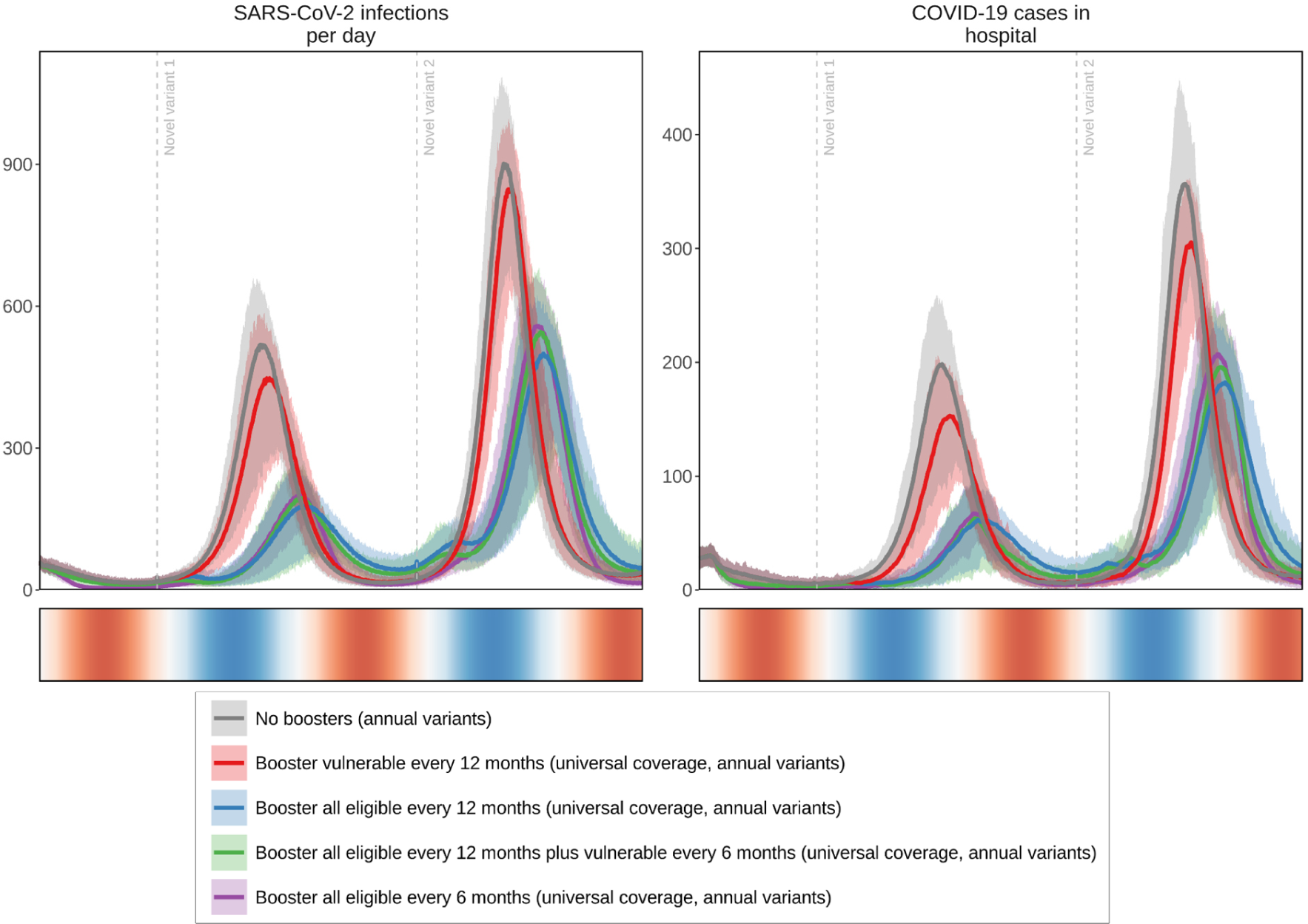
Projected daily impact on SARS-CoV-2 infections and COVID-19-related cases in hospital for just over a two-year period in a simulated population of 100,000 individuals. Those who were modelled as having previously received vaccine doses one and two, including 93% of those 60+ or with comorbidities, 90% of 50−59 year olds, 80% of 30−49 year olds, and 75% of 5−29 year olds, are eligible to receive first-generation COVID-19 vaccine boosters. Populations who received boosters every twelve or six months are aggregated into two groups, (1) those most vulnerable (60 years and older or persons with comorbidities) and (2) all eligible (five years and older). Each vaccine scenario curve is plotted alongside a baseline scenario curve whereby no vaccine booster doses were administered (grey curves). Booster dose coverage is 98% of those eligible (universal coverage). Novel SARS-CoV-2 variants with 25% more infectiousness and immune evading capacity than the previously dominant variant but with the same severity as the Omicron variant emerged annually (with initial emergence indicated with horizontal dashed lines). Shaded areas represent the stochastic uncertainty surrounding projections. Cumulative number and timing of vaccine doses are shown for each scenario. Seasonality is illustrated in the bottom row where red shading indicates the warmer spring and summer seasons, blue the cooler fall and winter seasons, and white the seasonal transition periods. This figure shows the same results as Figure S4A but with curves plotted within the same outcome panel.

**Figure S5A:**
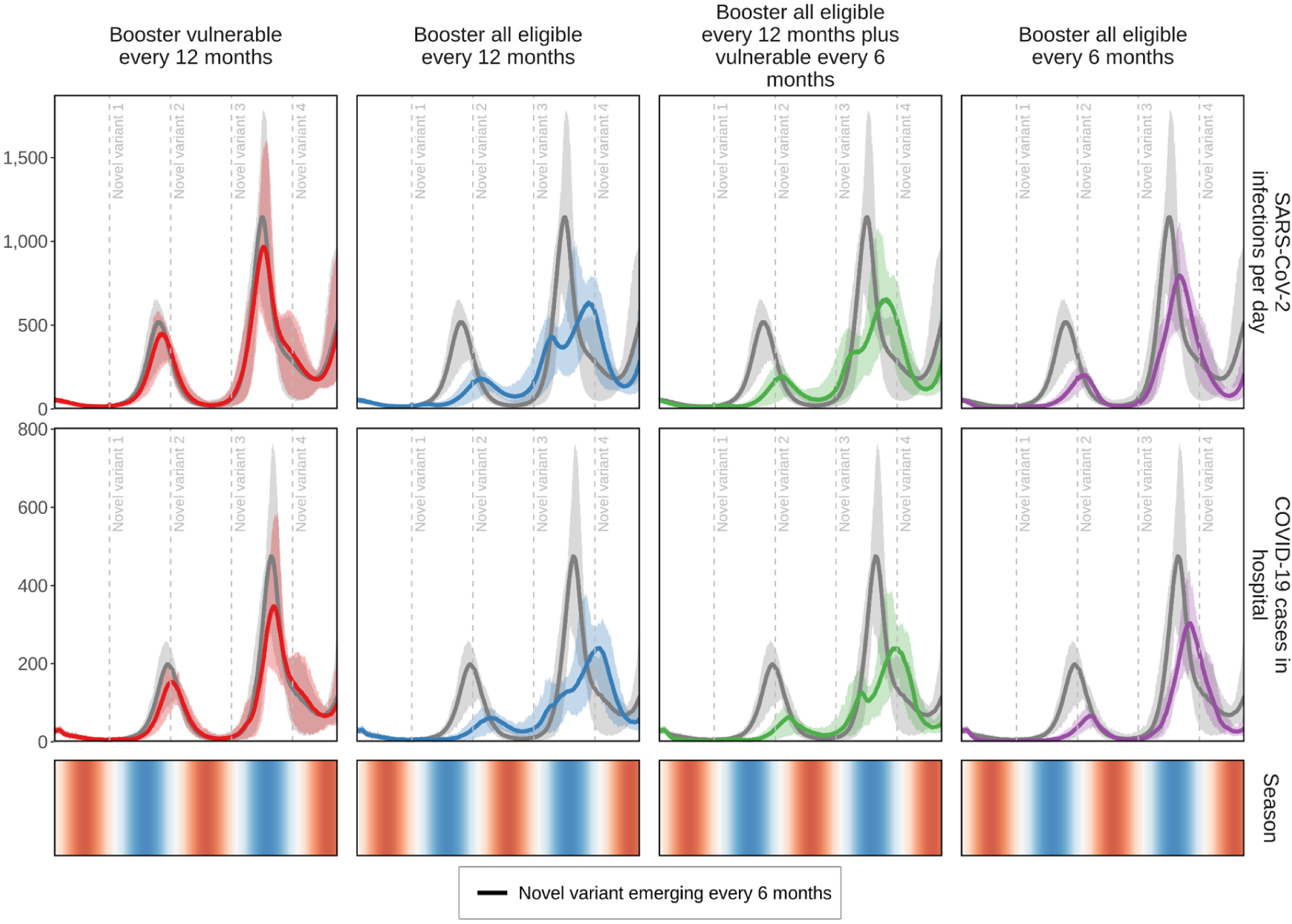
Projected daily impact on SARS-CoV-2 infections and COVID-19-related hospital admissions for just over a two-year period in a simulated population of 100,000 individuals. Those who were modelled as having previously received vaccine doses one and two, including 93% of those 60+ or with comorbidities, 90% of 50−59 year olds, 80% of 30−49 year olds, and 75% of 5−29 year olds, are eligible to receive first-generation COVID-19 Populations who received boosters every twelve or six months are aggregated into two groups, (1) those most vulnerable (60 years and older or persons with comorbidities) and (2) all eligible (those aged five years and older). Each vaccine scenario curve is plotted alongside a baseline scenario (grey curve) whereby no vaccine booster doses were administered (grey curves). Booster dose coverage is 98% of those eligible (universal coverage). Novel SARS-CoV-2 variants with 25% more infectiousness and immune evading capacity than the previously dominant variant but with the same severity as the Omicron variant emerged biannually (initial emergence indicated with horizontal dashed lines). Shaded areas represent the stochastic uncertainty surrounding projections. Cumulative number and timing of vaccine doses are shown for each scenario. Seasonality is illustrated in the bottom row where red shading indicates the warmer spring and summer seasons, blue the cooler fall and winter seasons, and white the seasonal transition periods. This figure is an extension of Figure 1 from the main text.

**Figure S5B:**
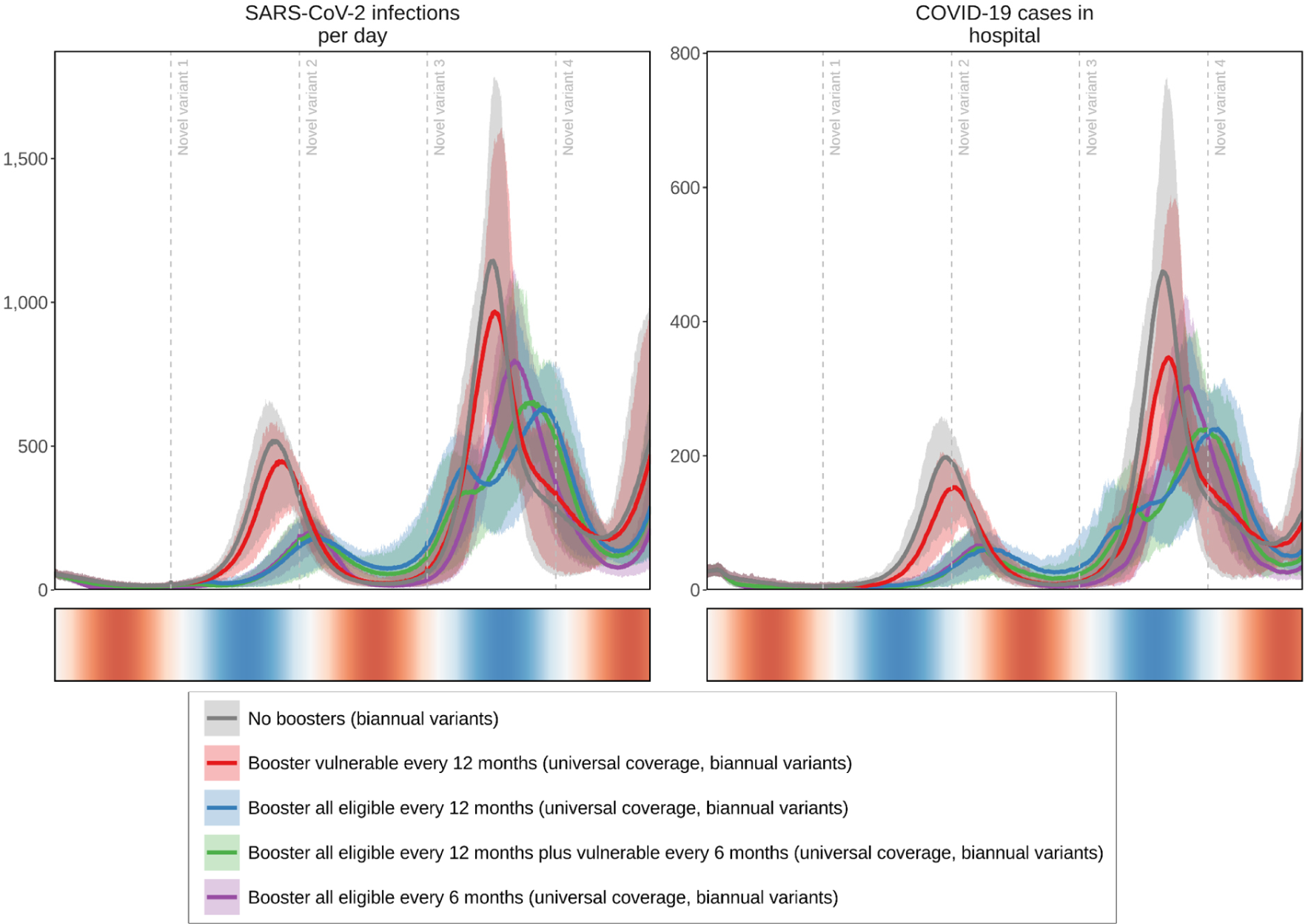
Projected daily impact on SARS-CoV-2 infections and COVID-19-related hospital admissions for just over a two-year period in a simulated population of 100,000 individuals. Those who were modelled as having previously received vaccine doses one and two, including 93% of those 60+ or with comorbidities, 90% of 50−59 year olds, 80% of 30−49 year olds, and 75% of 5−29 year olds, are eligible to receive first-generation COVID-19 Populations who received boosters every twelve or six months are aggregated into two groups, (1) those most vulnerable (60 years and older or persons with comorbidities) and (2) all eligible (five years and older). Each vaccine scenario curve is plotted alongside a baseline scenario curve whereby no vaccine booster doses were administered (grey curves). Booster dose coverage is 98% of those eligible (universal coverage). Novel SARS-CoV-2 variants with 25% more infectiousness and immune evading capacity than the previously dominant variant but with the same severity as the Omicron variant emerged biannually (initial emergence indicated with horizontal dashed lines). Shaded areas represent the stochastic uncertainty surrounding projections. Cumulative number and timing of vaccine doses are shown for each scenario. Seasonality is illustrated in the bottom row where red shading indicates the warmer spring and summer seasons, blue the cooler fall and winter seasons, and white the seasonal transition periods. This figure shows the same results as Figure S5A but with curves plotted within the same outcome panel.

**Figure S6:**
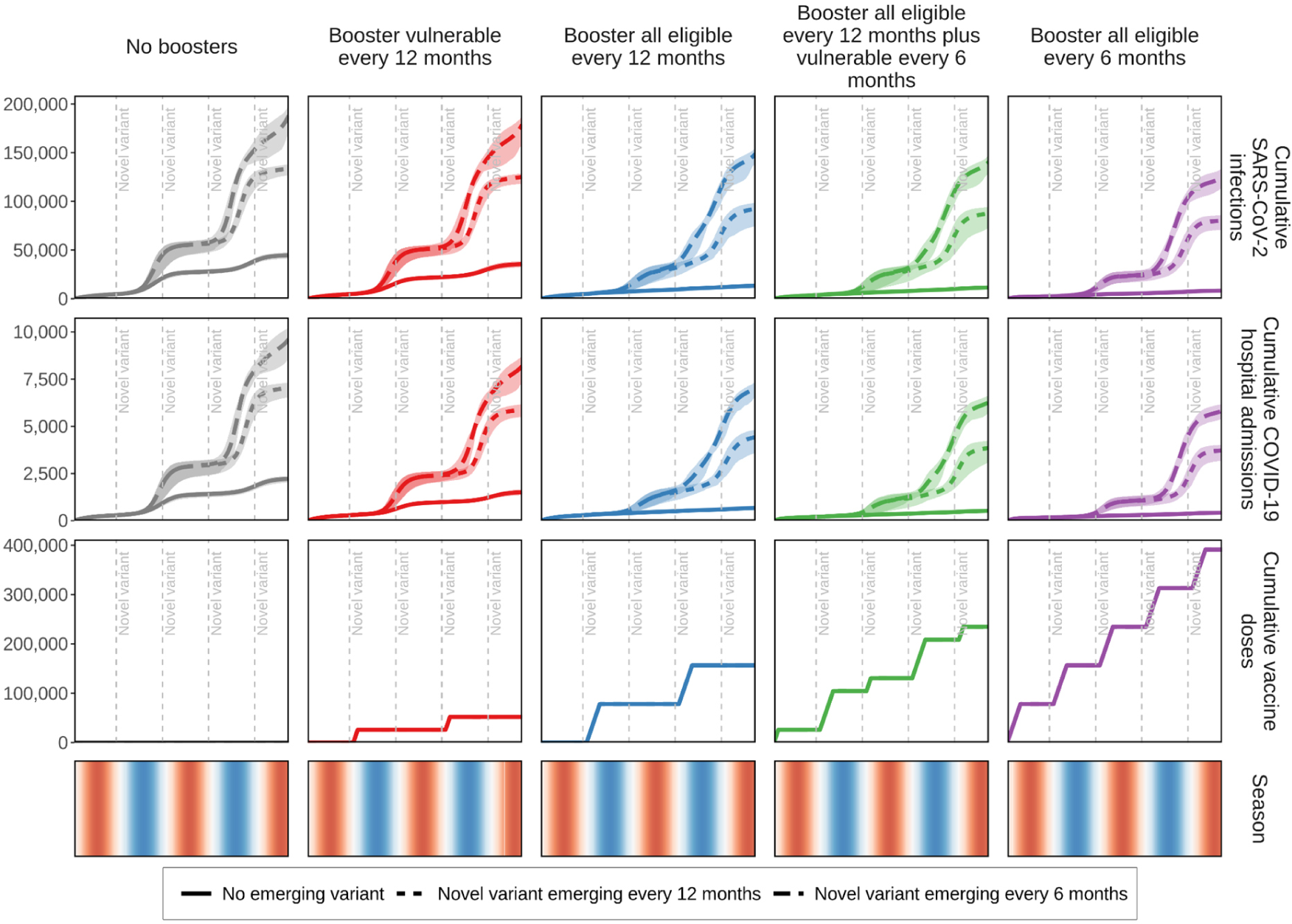
Cumulative projected impact on SARS-CoV-2 infections and COVID-19-related hospital admissions for just over a two-year period in a simulated population of 100,000 individuals. Those who were modelled as having previously received vaccine doses one and two, including 93% of those 60+ or with comorbidities, 90% of 50−59 year olds, 80% of 30−49 year olds, and 75% of 5−29 year olds, are eligible to receive first-generation COVID-19 We examined two main target groups who received boosters every twelve or six months, (1) those most vulnerable (60 years and older or persons with comorbidities), and (2) all eligible (five years and older). Booster dose coverage is 98% of those eligible (universal coverage). The impact of the baseline scenario (no boosters) with no vaccine boosters is represented by grey curves (left panels). It was simulated that there was either no novel emerging SARS-CoV-2 variant, assuming the same infectiousness and severity of the Omicron variant (solid curves); 25% more infectious and immune evading novel variants emerged every twelve months (short dashed curves); or every six months (long dashed curves) (timing of emerging variant indicated by initial emergence indicated with horizontal dashed lines). Shaded areas represent the stochastic uncertainty surrounding projections. Cumulative number and timing of vaccine doses are shown for each scenario. Seasonality is illustrated in the bottom row where red shading indicates the warmer spring and summer seasons, blue the cooler fall and winter seasons, and white the seasonal transition periods. This figure is an extension of Figure 2, right panels from the main text.

**Figure S7:**
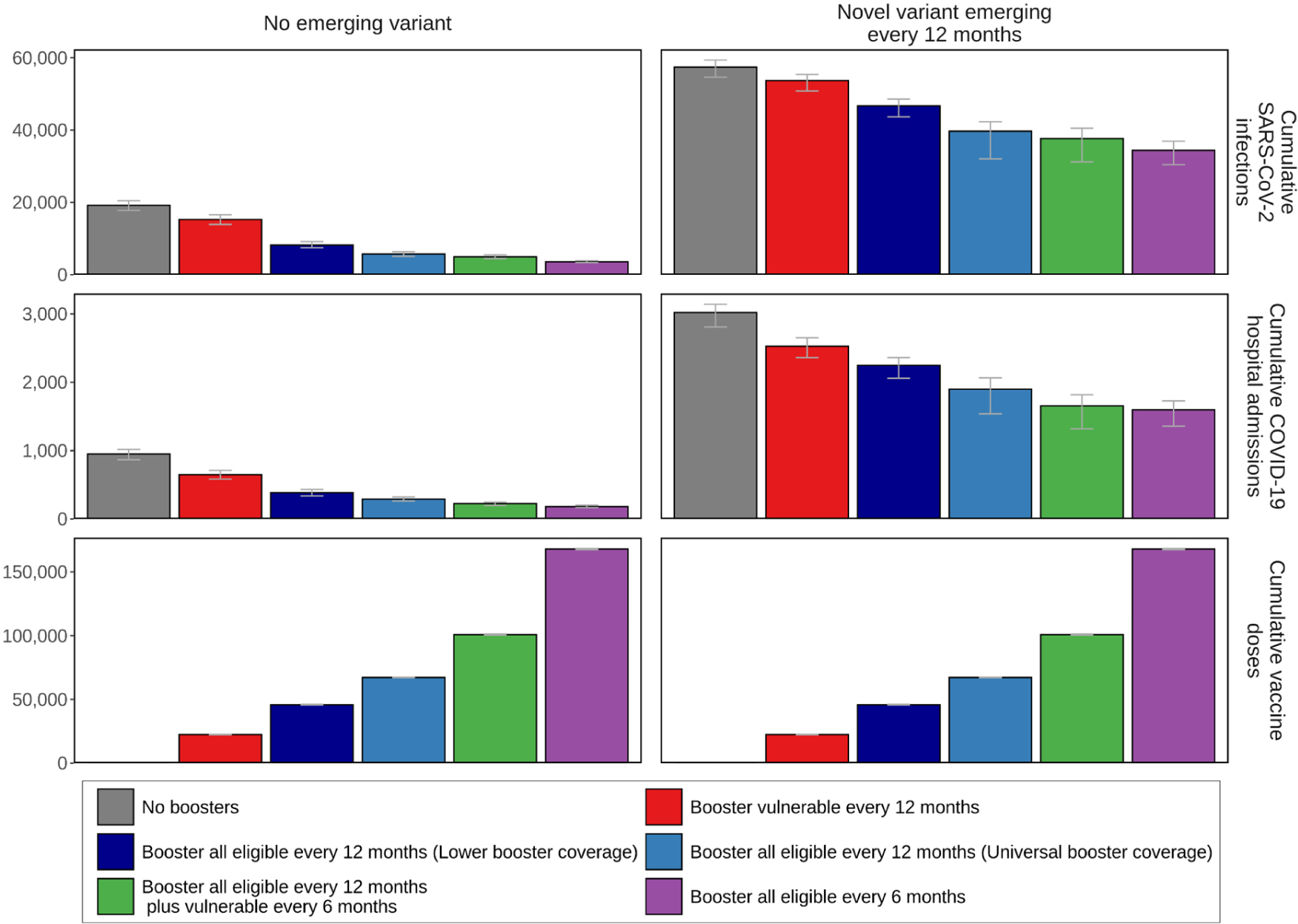
Cumulative projected impact on SARS-CoV-2 infections and COVID-19-related hospital admissions for just over a two-year period in a simulated population of 100,000 individuals. Those who were modelled as having previously received vaccine doses one and two, including 93% of those 60+ or with comorbidities, 90% of 50−59 year olds, 80% of 30−49 year olds, and 75% of 5−29 year olds, are eligible to receive first-generation COVID-19 Populations who received boosters every twelve or six months are aggregated into two main groups, (1) those most vulnerable (60 years and older or persons with comorbidities), and (2) all eligible (five years and older). Boosting those most vulnerable every six months includes boosting all those eligible every twelve months (green bars). Grey bars represent the impact of the baseline scenario with no vaccine boosters. Lower booster coverage was simulated as 85% coverage of those eligible to receive boosters who are most vulnerable (60 years and older or persons with comorbidities) and those 50−59 years of age, and 50% of those eligible aged 30−49 and 5−29 years. Universal booster coverage represents 98% of those eligible received booster doses. There was either no novel emerging SARS-CoV-2 variant, assuming the same infectiousness and severity of the Omicron variant (left panels), or novel variants with 25% more infectiousness and immune evading capacity emerged every twelve months (right panels). Error bars shown represent the stochastic uncertainty in model projections. This figure is a bar chart representation for cumulative impact curves from Figures 2 and 3 from the main text.

**Figure S8:**
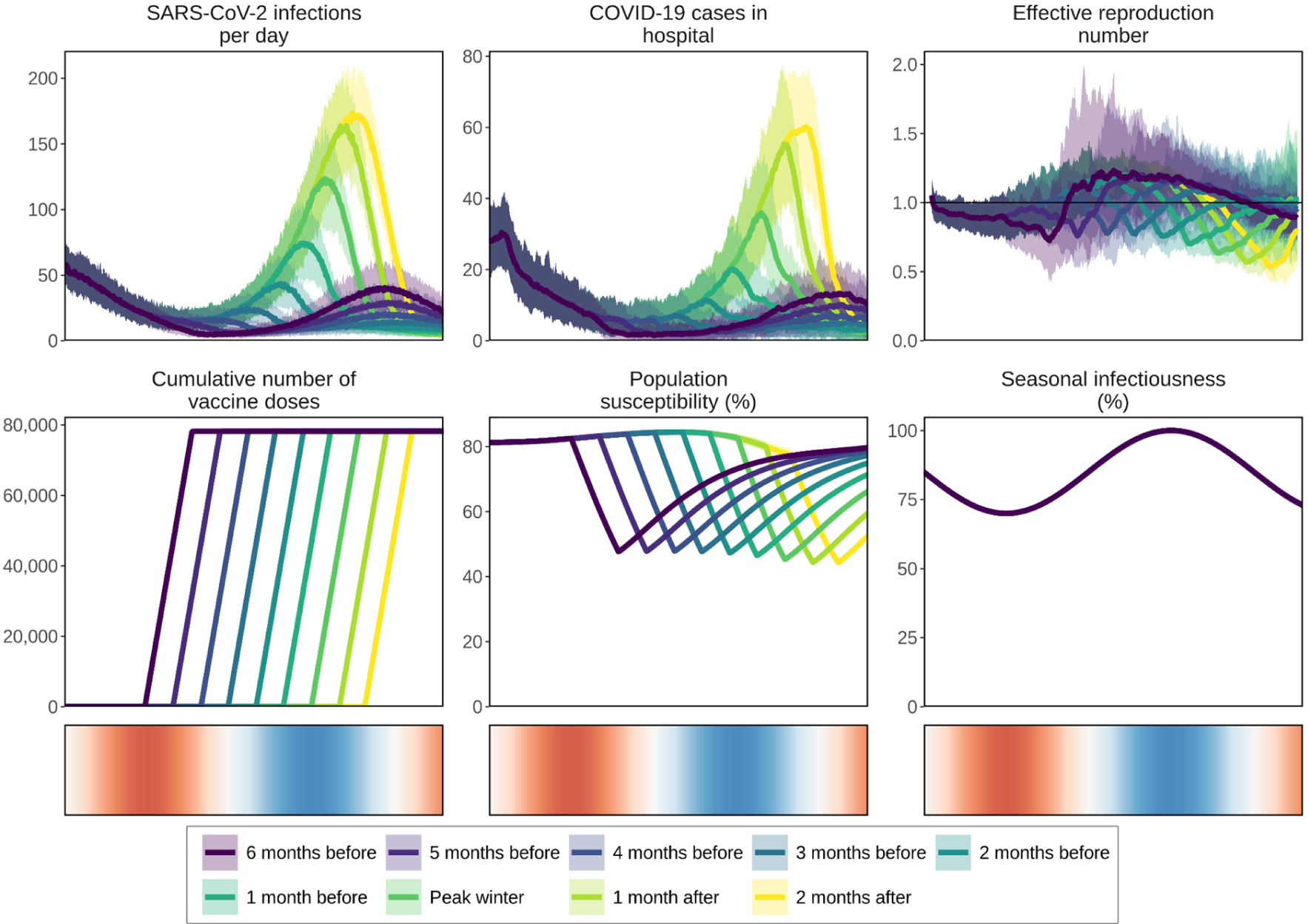
Projected daily impact on new COVID-19 infections, COVID-19-related hospital admissions, effective reproduction number, and population susceptibility of delivering vaccine boosters for all those eligible five years of age and older from either six months before to two months after peak winter temperatures assuming a 25% more infectious and immune evading novel variant emerges prior to the fall seasons. Shaded areas in the panels in the top row represent the stochastic uncertainty surrounding projections. Cumulative number of vaccine booster doses shown for each rollout start period. Seasonal infectiousness percentage plotted over time as a cosine function (bottom row, right panel) and illustrated in the bottom row where red shading indicates the warmer spring and summer seasons, blue the cooler fall and winter seasons, and white the seasonal transition periods.

**Figure S9:**
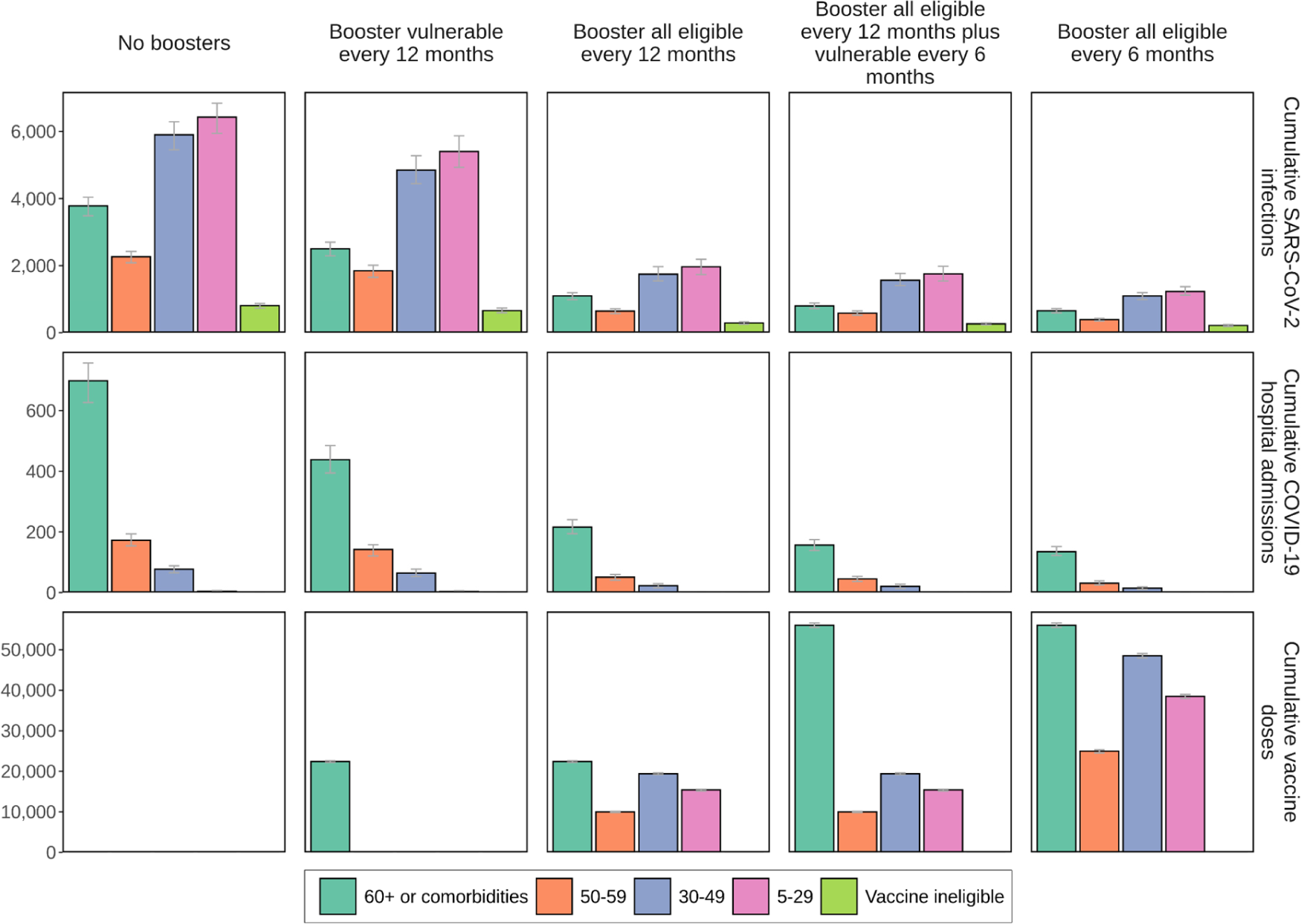
The cumulative projected impact of administering COVID-19 vaccine boosters to those most vulnerable or those eligible every twelve or six months. on SARS-CoV-2 infections and COVID-19- related hospital admissions by age and risk group over a two-year period in a simulated population of 100,000 individuals when new SARS-CoV-2 variants with 25% more infectiousness than the previously dominant variant but with the same severity as the Omicron variant emerged annually. Cumulative numbers of vaccine doses are shown in the bottom row. Error bars represent the stochastic uncertainty in model projections.

**Figure S10:**
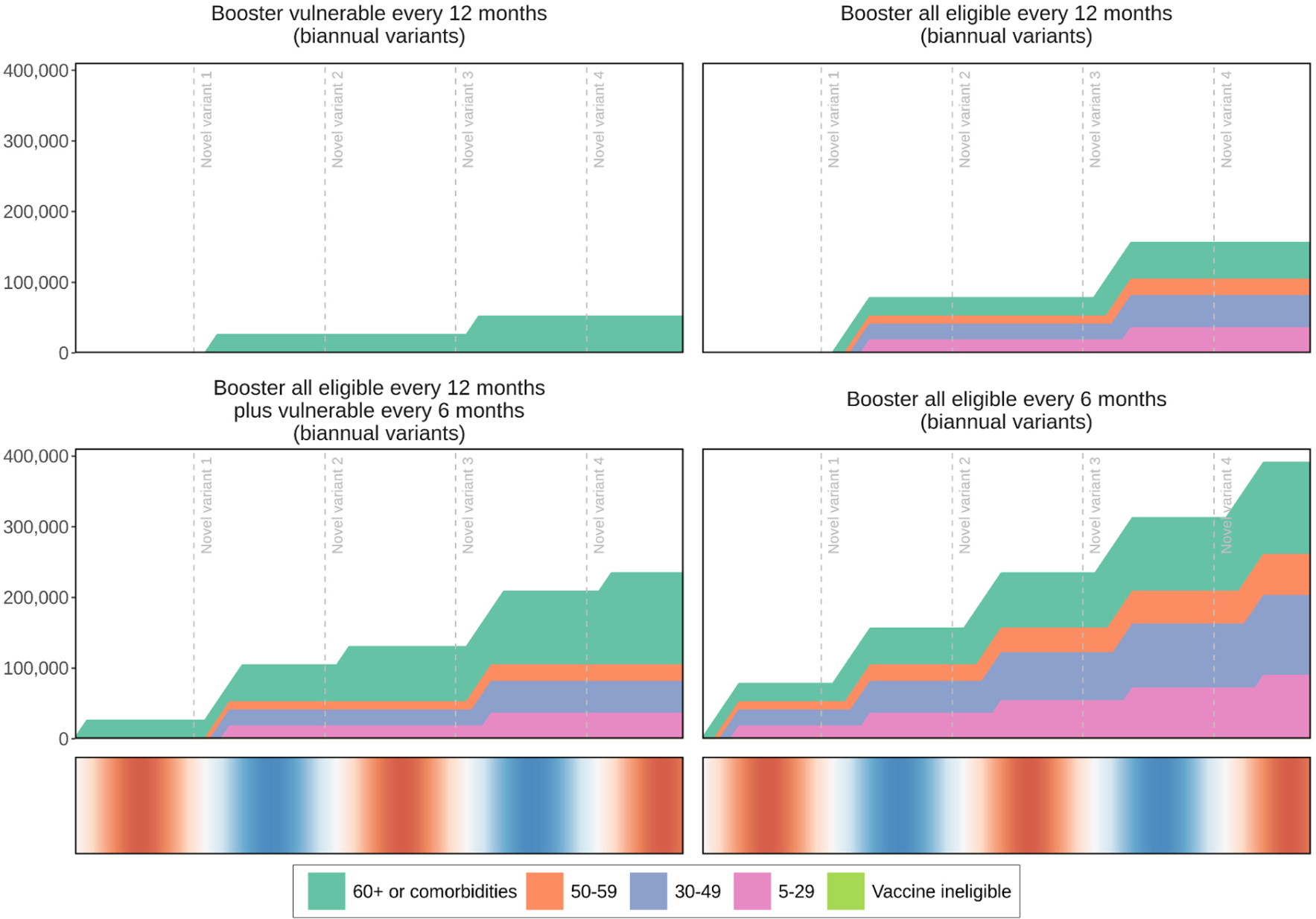
Cumulative COVID-19 vaccine booster dose administered by age and risk group for just over a two-year period in a simulated population of 100,000 individuals with those who previously received vaccine doses one and two are eligible to receive first-generation COVID-19 vaccine booster doses every twelve or six months. Those ineligible do not receive vaccine boosters (legend label corresponding to the light green shading). Boosting those most vulnerable every six months includes boosting all those eligible every twelve months (bottom row, left panel). Booster coverage was 98% of those who were eligible (universal coverage). Initial emergence indicated with horizontal dashed lines indicating where novel SARS-CoV-2 variants with a 25% more infectious and immune evading profile than the previously emerging variant were simulated to emerge every six months (the initial emergence of novel variants is indicated with horizontal dashed lines). Seasonality is illustrated in the bottom row where red shading indicates the warmer spring and summer seasons, blue the cooler fall and winter seasons, and white the seasonal transition periods.

**Figure S11:**
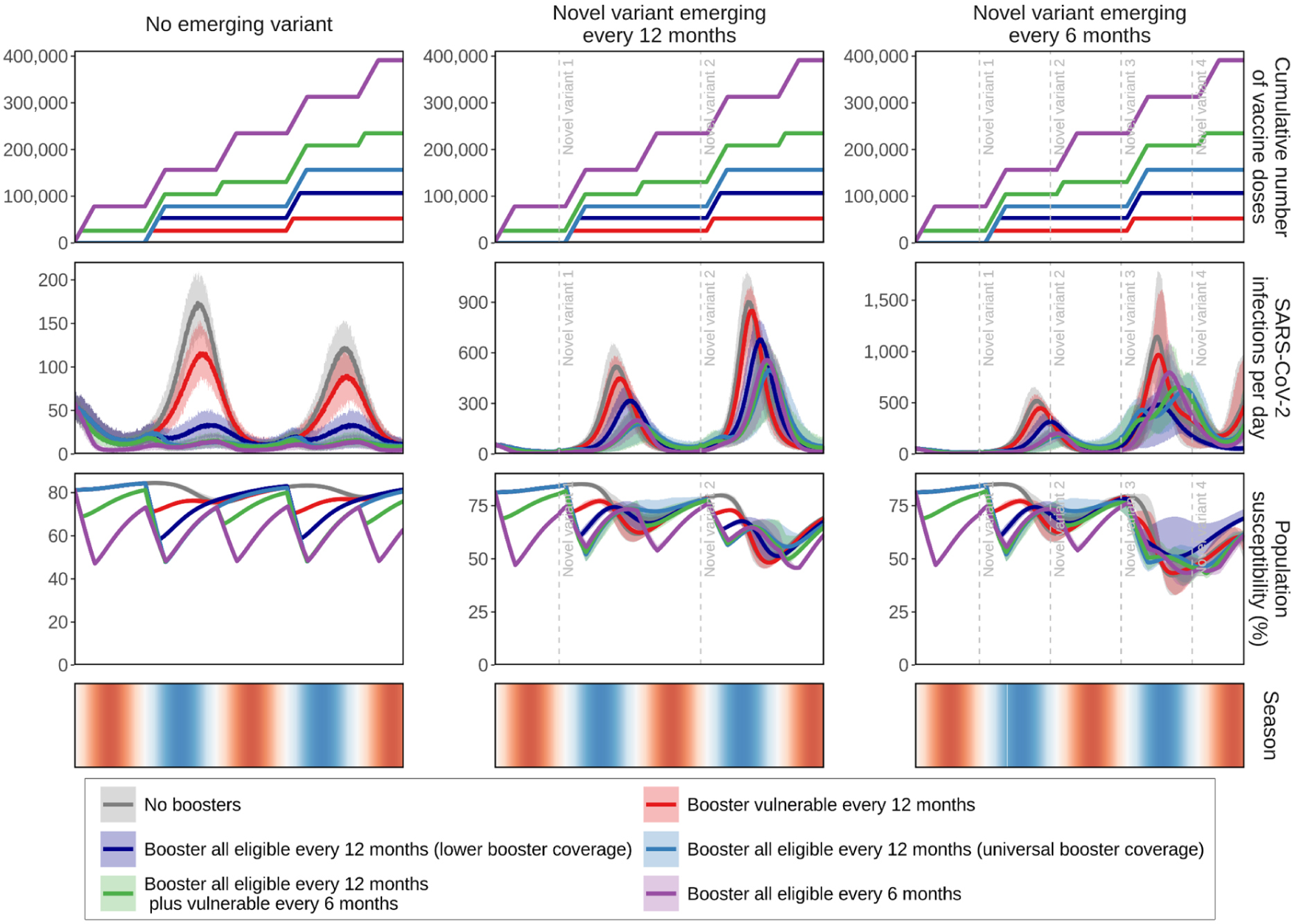
The cumulative number and timing of vaccine doses by scenario (top row, as shown in Figure 2 of the main text), projected daily impact of the various scenarios on SARS-CoV-2 infections (second row, corresponding to left panels from Figures S3B, S4B, and S5B), and daily percentage of population susceptibility by scenario (third row), in a simulated population of 100,000 individuals over a two-year period. People who previously received vaccine doses one and two are eligible to receive first-generation COVID-19 vaccine boosters. Those eligible received boosters every twelve or six months were aggregated into two groups, (1) those most vulnerable (60 years and older or persons with comorbidities) and (2) all eligible (five years and older). Boosting those most vulnerable every six months includes boosting all eligible people every twelve months. Grey curves represent the baseline scenario with no boosters modelled. Lower booster coverage (dark blue curve) was simulated as 85% coverage of those eligible 50 years of age and older and with comorbidities, and 50% of those eligible aged 5−49. Universal coverage (light blue curve) is 98% of those eligible to receive boosters. Scenarios were simulated as having (i) no novel emerging SARS-CoV-2 variant, assuming the same infectiousness and severity of the Omicron variant (left panels), (ii) novel variants with 25% more infectiousness and immune evading capacity emerging every twelve months (center panels), or (iii) emerging every six months (right panels) (initial emergence indicated with horizontal dashed lines). Shaded areas represent the stochastic uncertainty surrounding projections. Seasonality is illustrated in the bottom row where red shading indicates the warmer spring and summer seasons, blue the cooler fall and winter seasons, and white the seasonal transition periods.

**Figure S12A:**
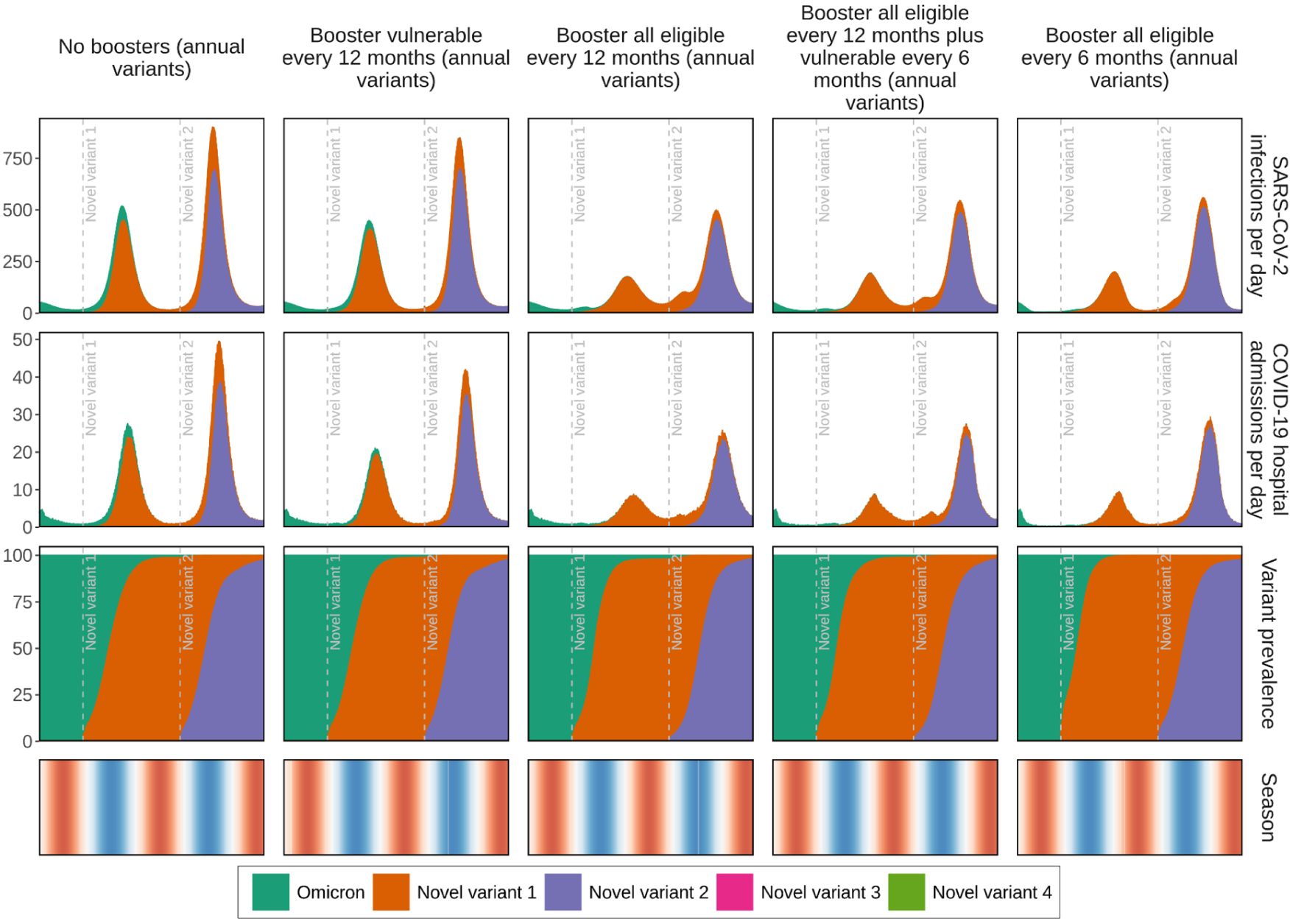
Projected impact of COVID-19 vaccine boosters on new daily SARS-CoV-2 infections and COVID-19-related hospital admissions by novel SARS-CoV-2 variant (stacked shaded areas under the curves) for just over a two-year period in a simulated population of 100,000 individuals. Those who previously received vaccine doses one and two were eligible to receive first-generation COVID-19 vaccine boosters. Populations who received boosters every twelve or six months are aggregated into two groups, (1) those most vulnerable (60 years and older or persons with comorbidities) and (2) all eligible (five years and older). Universal coverage of booster doses is 98% of those eligible. Novel variants with 25% more infectiousness and immune evading capacity but with the same severity as the Omicron variant emerged annually (initial emergence indicated with horizontal dashed lines, legend labels for novel variant 3 (pink) and novel variant 4 (lime green) do not apply in this figure). Prevalence by variant is plotted in the third row. Seasonality is illustrated in the bottom row where red shading indicates the warmer spring and summer seasons, blue the cooler fall and winter seasons, and white the seasonal transition periods.

**Figure S12B:**
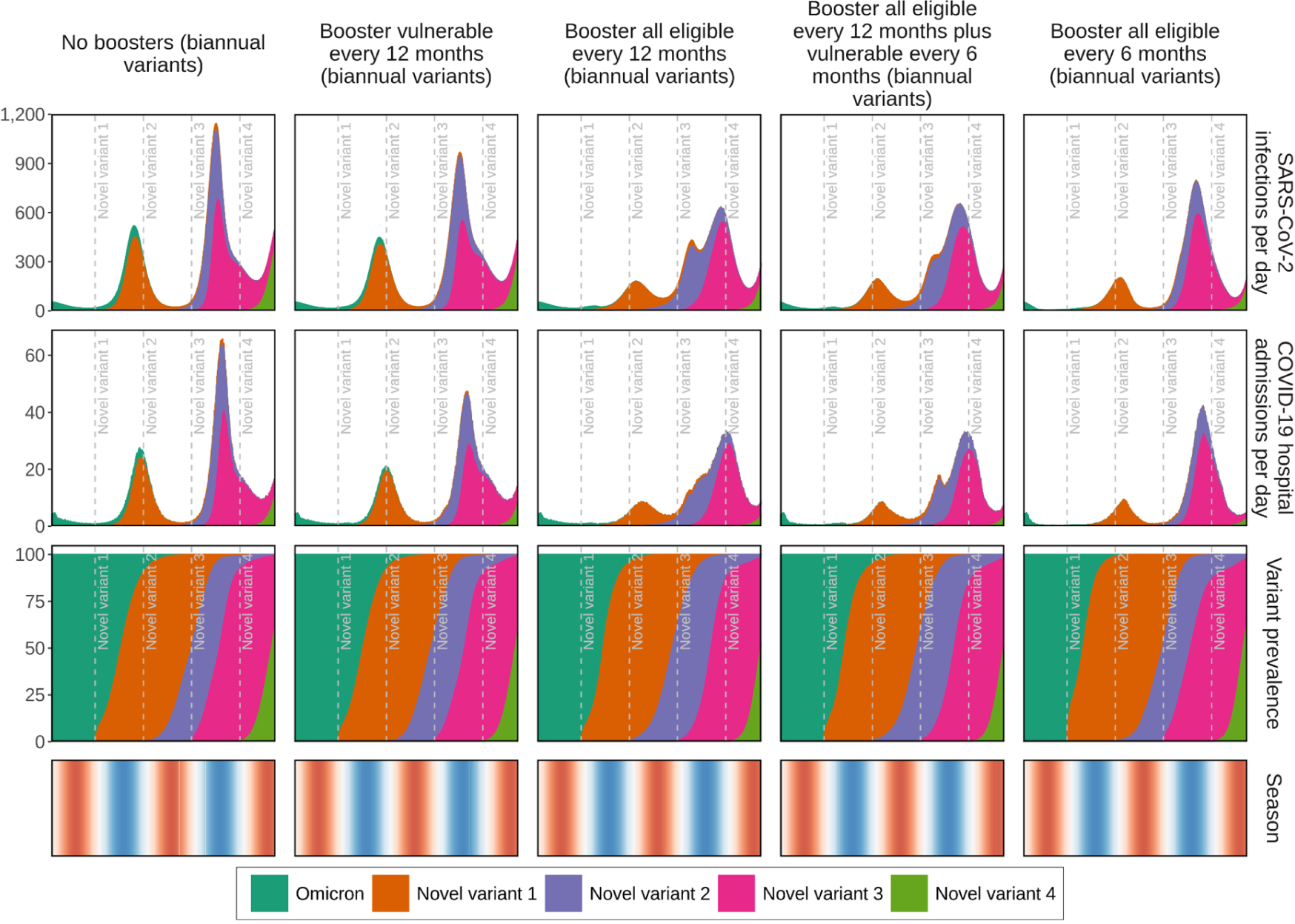
Projected impact of COVID-19 vaccine boosters on new daily SARS-CoV-2 infections and COVID-19-related hospital admissions by novel SARS-CoV-2 variant (stacked shaded areas under the curves) for just over a two-year period in a simulated population of 100,000 individuals. Those who previously received vaccine doses one and two were eligible to receive first-generation COVID-19 vaccine boosters. Populations who received boosters every twelve or six months are aggregated into two groups, (1) those most vulnerable (60 years and older or persons with comorbidities) and (2) all eligible (five years and older). Universal coverage of booster doses is 98% of those eligible. Novel variants with 25% more infectiousness and immune evading capacity emerged every six months (initial emergence indicated with initial emergence indicated with horizontal dashed lines). Prevalence by variant is plotted in the third row. Seasonality is illustrated in the bottom row where red shading indicates the warmer spring and summer seasons, blue the cooler fall and winter seasons, and white the seasonal transition periods.

**Figure S13A:**
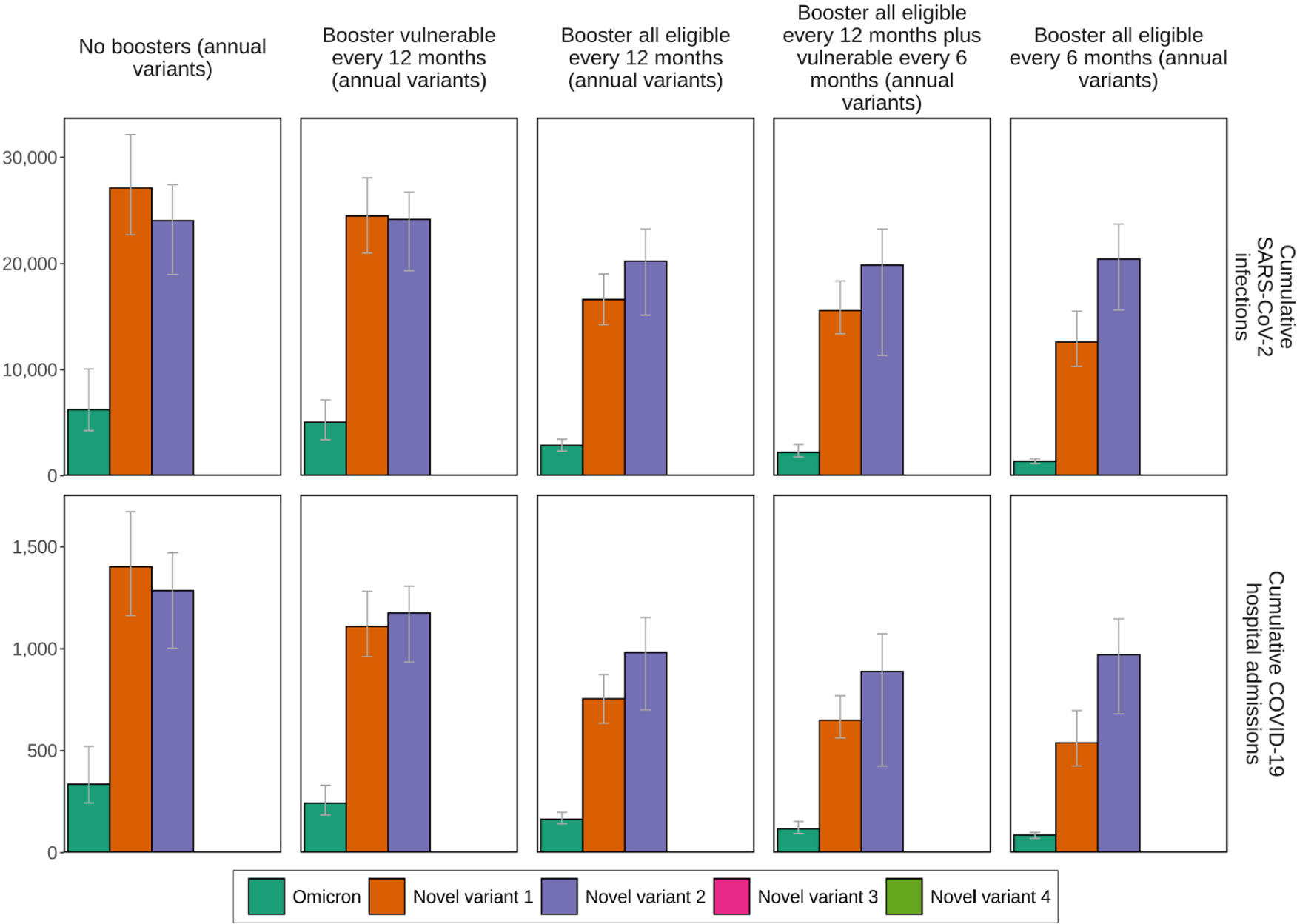
Cumulative projected impact of COVID-19 vaccine boosters on SARS-CoV-2 infections and COVID-19-related hospital admissions by variant when new SARS-CoV-2 variants with 25% more infectiousness and immune evading capacity but with the same severity as the Omicron variant emerged annually (legend labels for novel variant 3 (pink) and novel variant 4 (lime green) do not apply in this figure). This was simulated for just over a two-year period in a population of 100,000 individuals. Those who previously received vaccine doses one and two were eligible to receive first-generation COVID-19 vaccine boosters. Populations who received boosters every twelve or six months are aggregated into two main groups, (1) those most vulnerable (60 years and older or persons with comorbidities), and (2) all eligible (five years and older). Boosting those most vulnerable every six months includes boosting all those eligible every twelve months. Universal booster coverage represents 98% of those eligible received booster doses. Error bars shown represent the stochastic uncertainty in model projections.

**Figure S13B:**
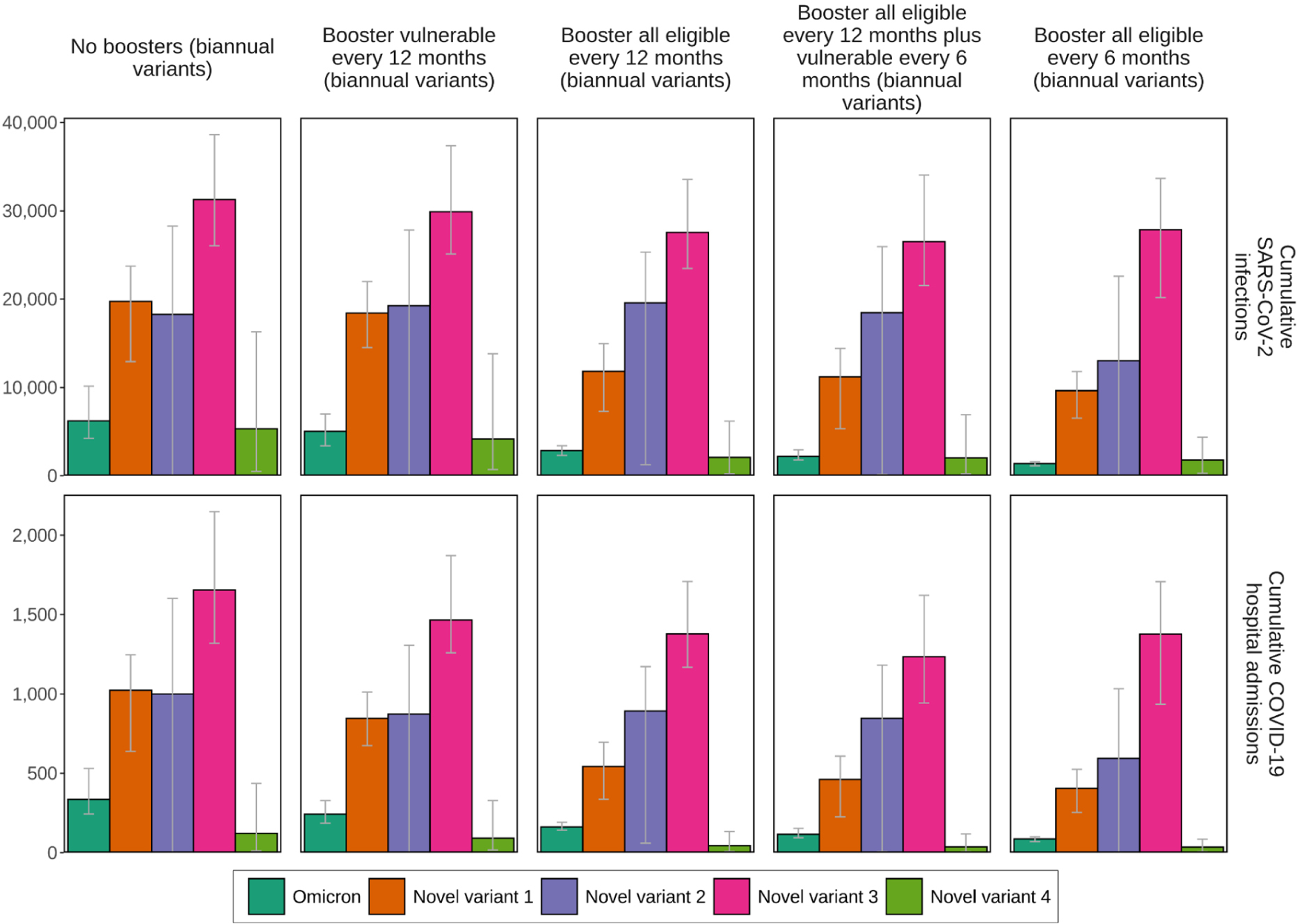
Cumulative projected impact of COVID-19 vaccine boosters on SARS-CoV-2 infections and COVID-19-related hospital admissions by variant when novel SARS-CoV-2 variants with 25% more infectiousness and immune evading capacity emerged every six months. This was simulated for just over a two-year period in a population of 100,000 individuals. Those who previously received vaccine doses one and two were eligible to receive first-generation COVID-19 vaccine boosters. Populations who received boosters every twelve or six months are aggregated into two main groups, (1) those most vulnerable (60 years and older or persons with comorbidities), and (2) all eligible (five years and older). Boosting those most vulnerable every six months includes boosting all those eligible every twelve months. Universal booster coverage represents 98% of those eligible received booster doses. Error bars shown represent the stochastic uncertainty in model projections. Novel variant four only emerges at the end of the simulation period; therefore, its full impact is not yet captured.

**Figure S14A:**
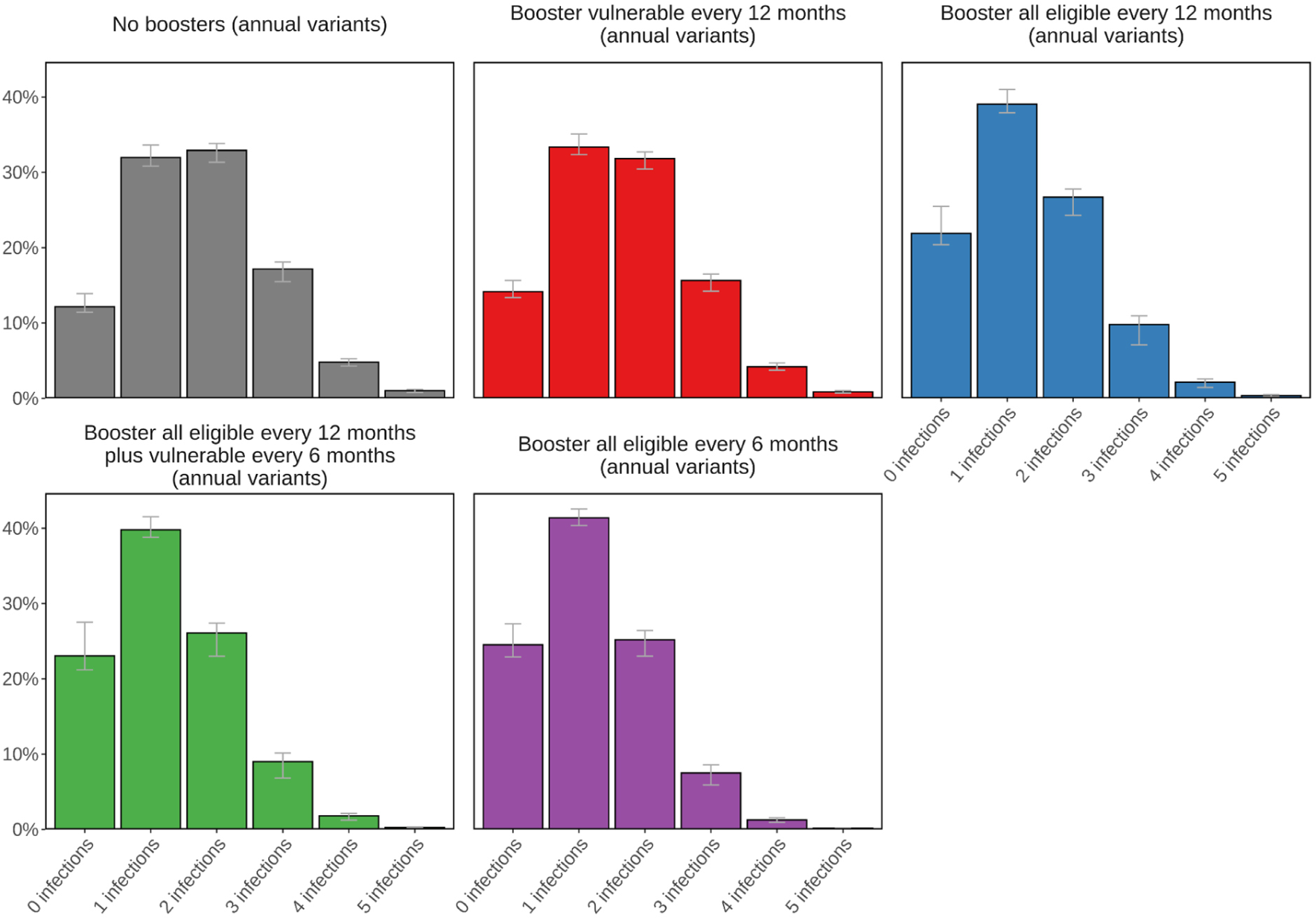
Cumulative projected impact of COVID-19 vaccine boosters on the percentage of SARS-CoV-2 infections (and re-infections) when novel SARS-CoV-2 variants with 25% more infectiousness and immune evading capacity emerged annually. This was simulated for just over a two-year period in a population of 100,000 individuals. Those who previously received vaccine doses one and two were eligible to receive first-generation COVID-19 vaccine boosters. Populations who received boosters every twelve or six months are aggregated into two main groups, (1) those most vulnerable (60 years and older or persons with comorbidities), and all eligible (five years and older). Boosting those most vulnerable every six months includes boosting all those eligible every twelve months. Grey bars represent infections from the baseline scenario with no vaccine boosters. Universal booster coverage represents 98% of those eligible received booster doses. Error bars shown represent the stochastic uncertainty in model projections.

**Figure S14B:**
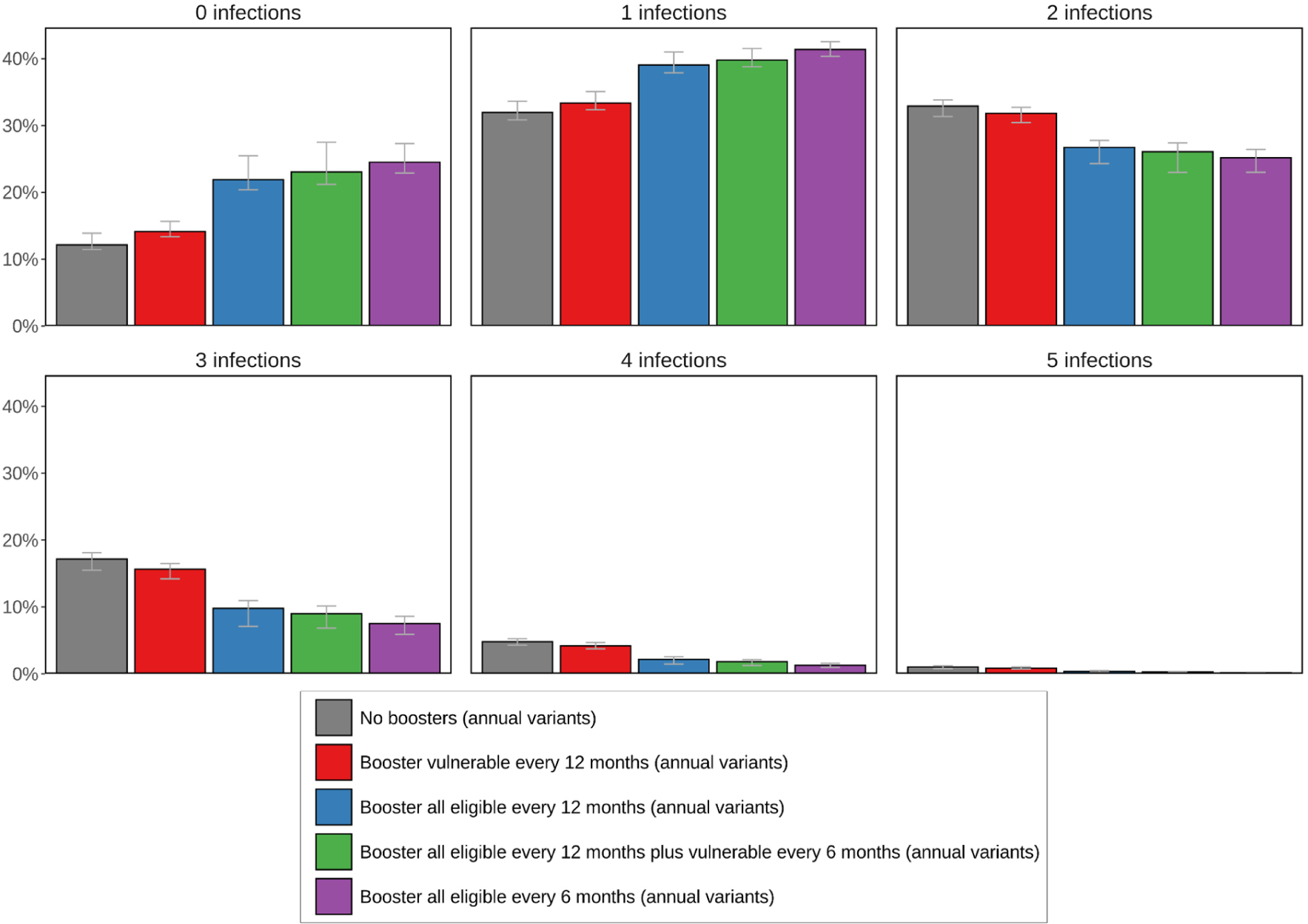
Cumulative projected impact of COVID-19 vaccine boosters on the percentage of SARS-CoV-2 infections (and re-infections) grouped by number of infections when novel SARS-CoV-2 variants with 25% more infectiousness and immune evading capacity emerged annually. This was simulated for just over a two-year period in a population of 100,000 individuals. Those who previously received vaccine doses one and two were eligible to receive first-generation COVID-19 vaccine boosters. Populations who received boosters every twelve or six months are aggregated into two main groups, (1) those most vulnerable (60 years and older or persons with comorbidities), and (2) all eligible (five years and older). Boosting those most vulnerable every six months includes boosting all those eligible every twelve months. Grey bars represent infections from the baseline scenario with no vaccine boosters. Universal booster coverage represents 98% of those eligible received booster doses. Error bars shown represent the stochastic uncertainty in model projections.

**Figure S15:**
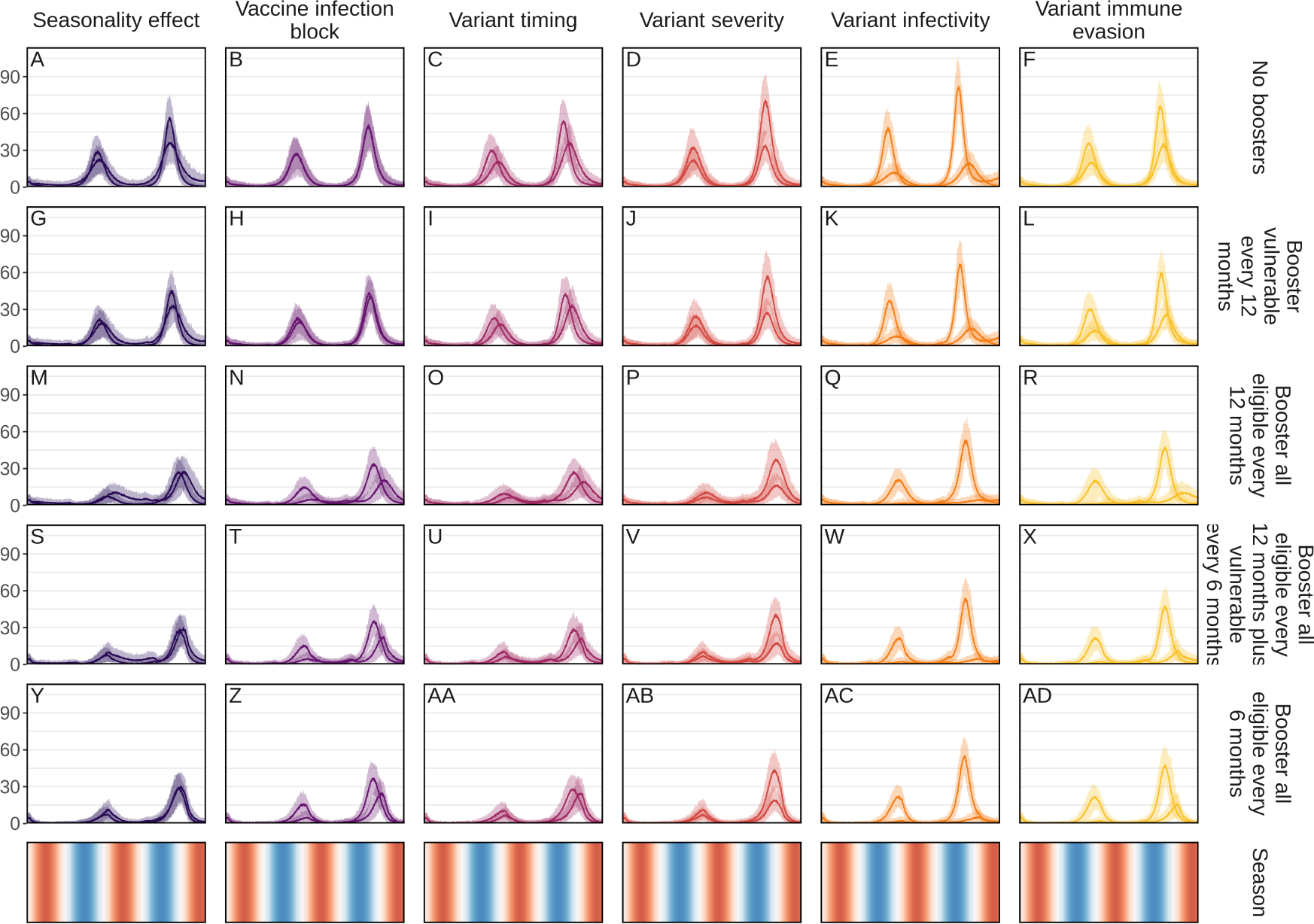
Sensitivity analysis showing the projected impact on COVID-19-related hospital admissions averted over a two-year period from modifying seasonality effect, vaccine infection blocking, variant timing, variant severity, variant infectivity, and variant immune evasion with the two curves in each panel representing the lower and upper bound values for each parameter as listed in Table 1 of the main text. This time series was simulated for just over a two-year period in a population of 100,000 individuals. Those who previously received vaccine doses one and two were eligible to receive first-generation COVID-19 vaccine boosters. Populations who received boosters every twelve or six months are aggregated into two main groups, (1) those most vulnerable (60 years and older or persons with comorbidities), and (2) all eligible (five years and older). Boosting those most vulnerable every six months includes boosting all those eligible every twelve months. No boosters (top row) represents the baseline scenario with no vaccine boosters administered. Seasonality is illustrated in the bottom row where red shading indicates the warmer spring and summer seasons, blue the cooler fall and winter seasons, and white the seasonal transition periods.

**Figure S16A:**
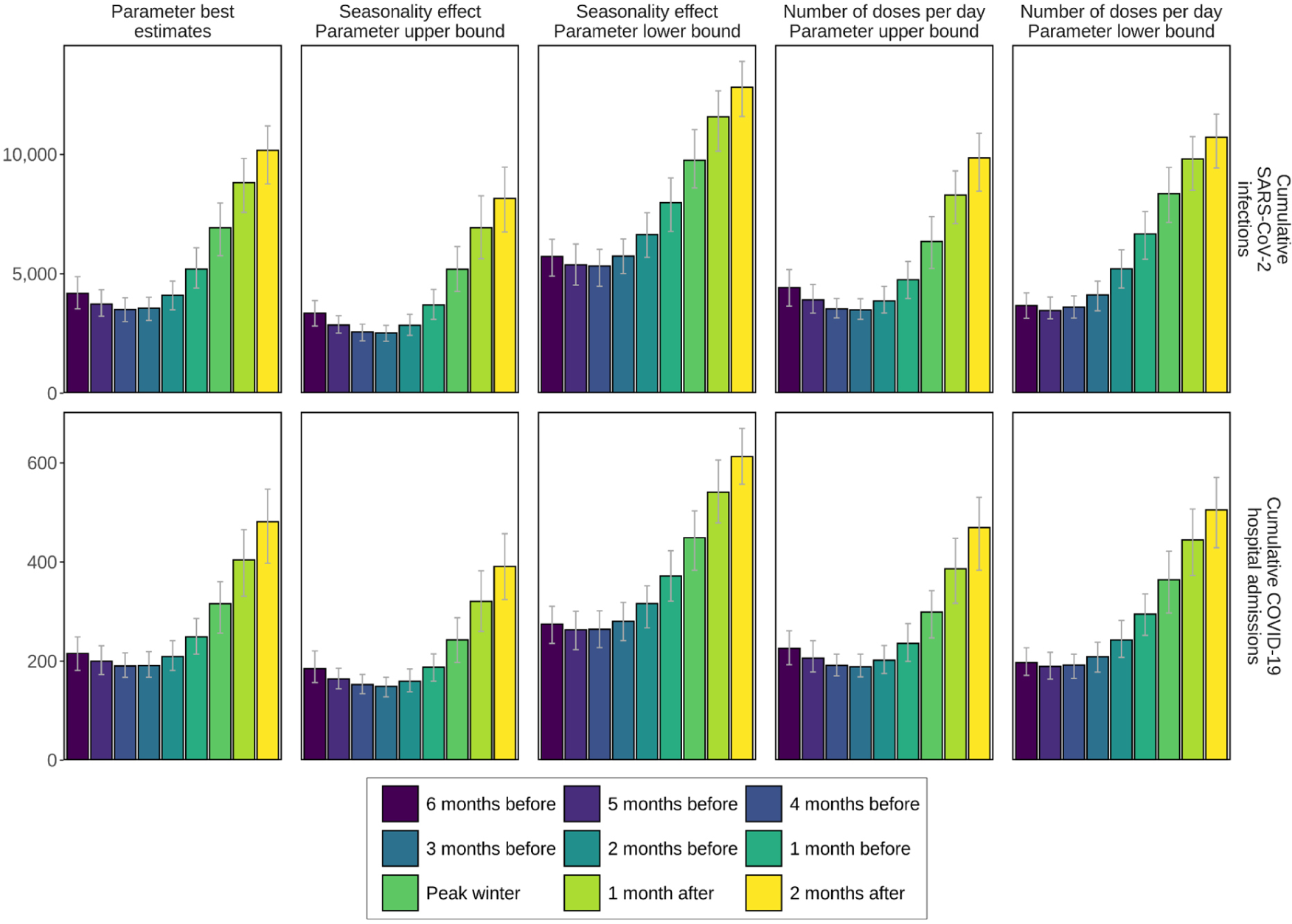
Impact of varying vaccine booster dose timing from six months before to two month after peak winter on the cumulative SARS-CoV-2 infections and COVID-19-related hospital admissions over a two-year period.

**Figure S16B:**
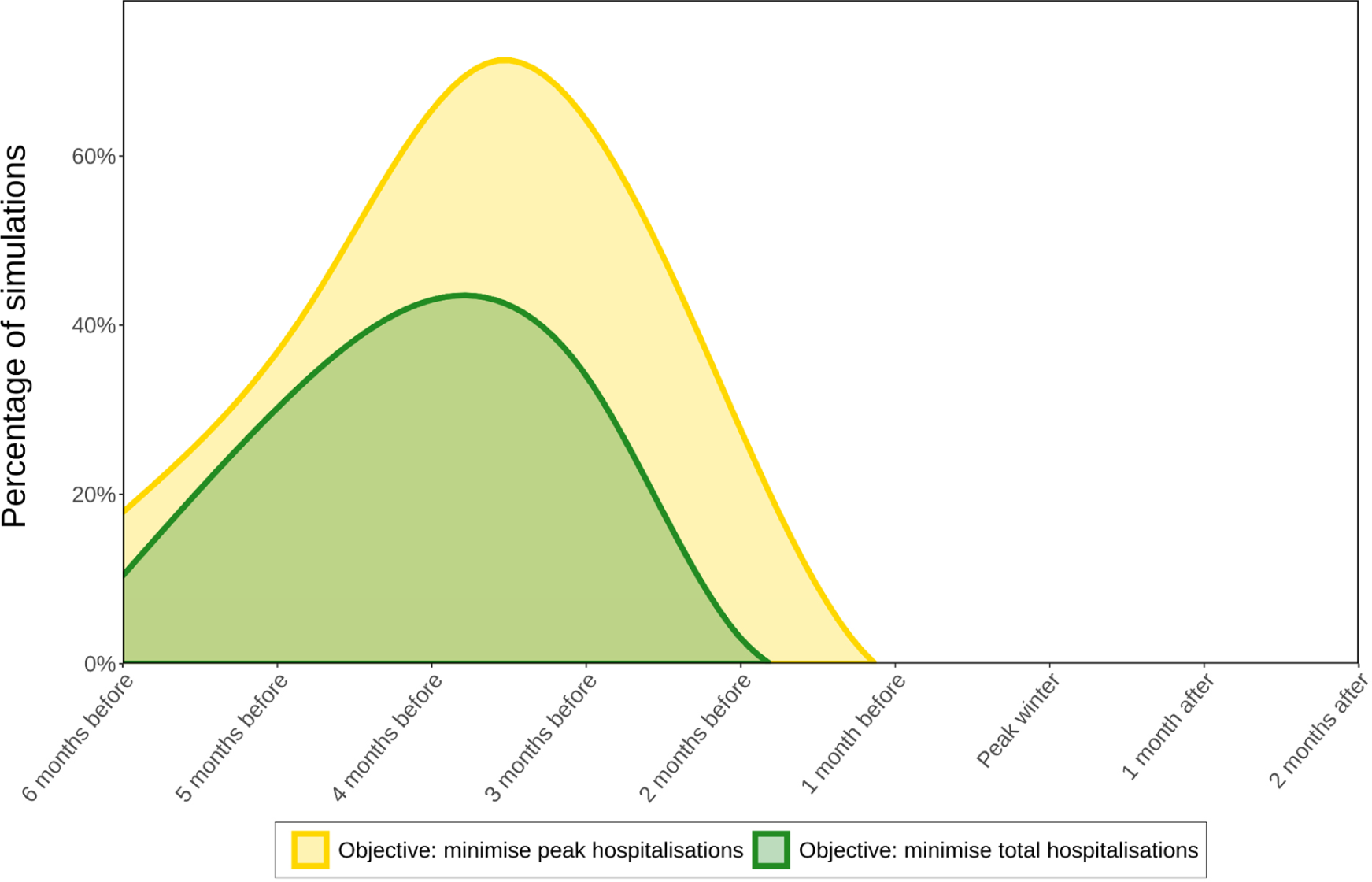
Percentage of simulations to optimise the start of COVID-19 vaccine booster roll-out considering parameter and stochastic uncertainty to either minimise peak hospitalisations (yellow area curve) or total hospitalisations (green area curve) from six months before to two months after peak winter.

**Figure S17:**
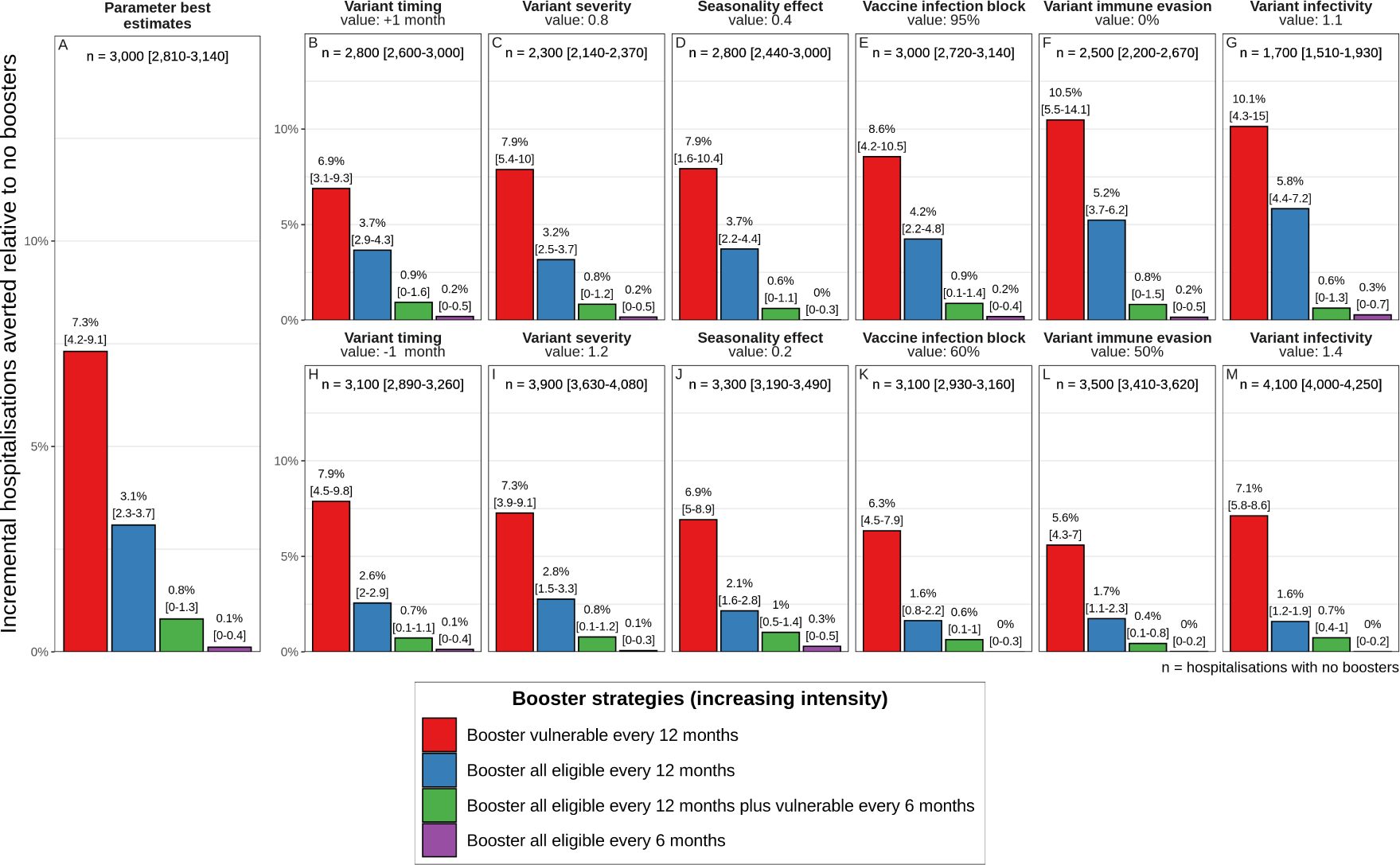
Percentage of COVID-19-related hospitalisations averted over a two-year period per 100,000 COVID-19 vaccine booster doses administered relative to no boosters given to those who received the first two doses and are most vulnerable (60 years and older or persons with comorbidities) or all eligible (five years and older) every twelve or six months, alongside varying the variant timing, variant severity, seasonality, vaccine infection blocking, variant immune evasion, and variant infectivity from best estimate values as listed in Table 1. Uncertainty bounds are shown in brackets above each bar. The numbers of hospitalisations with no boosters (n) with uncertainty bounds are shown at the top of each panel.

